# Association between saturated fat intake and low-density lipoprotein cholesterol across the genetic spectrum: Results from the Women’s Health Initiative

**DOI:** 10.64898/2026.06.29.26356854

**Authors:** Alexa Barad, Victor Ritter, Matthew Nudy, Linda Van Horn, Matthew A. Allison, Cassandra N. Spracklen, Longjian Liu, Su Yon Jung, JoAnn E. Manson, Themistocles L. Assimes, Marcia L. Stefanick, Shoa L. Clarke

## Abstract

**Background:** Elevated low-density lipoprotein cholesterol (LDL-C) is a causal risk factor for atherosclerotic cardiovascular disease (ASCVD). Guidelines recommend reducing saturated fat intake to lower LDL-C. However, the response to saturated fat in the diet is highly heterogeneous. Genetic factors may contribute to individual differences in response to saturated fat.

**Objectives:** We aimed to examine whether genetic propensity for higher LDL-C modifies the association of saturated fat intake with LDL-C and incident ASCVD.

**Methods:** We studied 20,940 genotyped postmenopausal women from the Women’s Health Initiative. Exposures included saturated fat intake (percentage of total calories) derived from food frequency questionnaires and a genome-wide polygenic score for LDL-C (PGS_LDL_). The primary outcome was LDL-C. The secondary outcome was incident ASCVD. Associations were assessed using multivariable linear and Cox regressions. Effect modification was evaluated using interaction terms and restricted cubic spline analyses. Models were adjusted for demographic, anthropometric, lifestyle, clinical, genetic, and technical covariates.

**Results:** The median LDL-C at baseline for participants with PGS_LDL_ below and above the median was 135 mg/dL [Q1: 114, Q3: 160] and 162 mg/dL [137, 188], respectively. Saturated fat intake was positively associated with LDL-C in the high PGS_LDL_ group, but the association attenuated in the low PGS_LDL_ group (*P*-interaction for LDL-C = 0.01). Spline analysis revealed a non-linear interaction between PGS_LDL_ and saturated fat, with modifying effects emerging at higher PGS_LDL_. Compared to individuals with low PGS_LDL_ and low saturated fat intake, only those with both high PGS_LDL_ and high saturated fat intake had increased risk for ASCVD in an adjusted analysis (HR 1.45, 95% CI 1.20-1.75). This association remained significant after further adjustment for baseline untreated LDL-C (HR 1.29, 95% CI 1.06-1.56), and evidence of an additive interaction was observed (relative excess risk due to interaction 0.37, 95% CI 0.06-0.67]).

**Conclusions:** These findings suggest that the association between saturated fat intake and LDL-C and subsequent ASCVD risk may be stronger for individuals with a genetic propensity towards high LDL-C.

## Introduction

Elevated low-density lipoprotein cholesterol (LDL-C) is the third largest contributor towards global cardiovascular morbidity and mortality.^1^ A healthy diet is central to controlling LDL-C, and clinical trials have firmly established saturated fat as a key macronutrient that drives elevation of LDL-C.^2,3^ Hence, both professional societies and the U.S. federal government have consistently recommended limiting saturated fat intake to less than 10% of total calories.^4–6^ Nonetheless, controversy regarding saturated fat intake as a risk factor for cardiovascular disease has persisted,^7,8^ and two thirds of U.S. adults exceed the recommended intake limit.^9^ This controversy may stem, in part, from the complex relationship between saturated fat and cardiovascular disease risk, which is influenced by several factors, including the nutrient replacing saturated fat^10^ and the food matrix in which it is consumed.^11^ Another important contributor to this controversy is the considerable heterogeneity in LDL-C responses to saturated fat intake,^12^ which may also contribute to uncertainty among patients. Indeed, the average effects observed in dietary intervention studies obscure the substantial inter-individual variability in LDL-C responses to dietary changes.^13–15^ Understanding the determinants of such variability is a critical step towards the development of more refined nutrition recommendations.

Genetic factors have long been hypothesized to influence the effects of diet on health. Several studies have implicated specific genetic variants at various lipid-related loci in modifying the effects of dietary factors on LDL-C.^16–23^ The breadth of these examples raises the possibility that overall genetic propensity towards higher or lower LDL-C may itself be a modifier of the effects of diet on lipids. If so, then contemporary polygenic scores could offer a powerful means to delineate such effects across the full spectrum of genetic backgrounds. A recent small study showed that low-carbohydrate, high total fat diets were associated with higher LDL-C compared to other diets among individuals in the top 20% of polygenic risk for hypercholesterolemia.^24^ Others have found that a “diet-response” polygenic score, derived using total fat intake, modestly predicted the effects of a low-fat diet intervention.^25^ These prior observations motivate the need for larger studies to examine the specific effects of saturated fat on LDL-C and cardiovascular outcomes across the full spectrum of genetic propensity.

Here, we use a validated genome-wide multi-ancestry polygenic score for LDL-C to capture the full distribution of polygenic propensity for higher or lower LDL-C.^26^ We examine whether polygenic risk modifies the association between saturated fat intake and LDL-C, as well as incident atherosclerotic cardiovascular disease (ASCVD), and we evaluate the shapes of these relationships. To do so, we analyzed >20,000 genotyped participants of the Women’s Health Initiative (WHI) with extensive dietary assessments, LDL-C measurements, and follow-up for incident ASCVD.

## Methods

### Study Cohort

The WHI study design and methods have been previously described.^27,28^ In brief, the WHI enrolled 161,808 postmenopausal women between the ages of 50 and 79 years from 40 clinical centers across the United States between 1993 and 1998. Participants were enrolled into the clinical trial (CT) component, which consisted of three randomized controlled trials (n = 68,132), or the observational study (OS) component (n = 93,676). At the end of the first phase of the WHI (2004-2005), women were invited to participate in two extension studies between 2005-2010 (Extension 1) and 2010-2015 (Extension 2). The WHI was approved by the institutional review boards at the clinical coordinating center and all 40 participating centers, and written informed consent was obtained from all participants.

A subset of women from both the CT and OS components underwent genotyping as part of nine ancillary studies (n = 42,400) and were considered for inclusion in this study. Of these, we excluded individuals who were related at the second-degree level or closer (n = 634), those genotyped using a lower-quality array (n = 153; two of the nine ancillary studies), and those whose genetically inferred ancestry fell outside the four largest genetically inferred ancestry groups (European [EUR], African [AFR], Admixed American [AMR], East Asian [EAS]). Among the remaining participants, 24,732 had baseline LDL-C measurements. Of these, we further excluded participants with missing or implausible (<600 or > 5000 kcal/day) baseline food frequency questionnaire (FFQ) data (n = 1,214), or with missing covariate data (n = 2,578). This resulted in a final analytic sample of 20,940 women for the primary cross-sectional LDL-C analyses. For the prospective analyses of incident ASCVD, we excluded women with prevalent ASCVD at enrollment (n = 1,457), which was defined as having at least one self-reported prior cardiovascular event or procedure, including history of myocardial infraction, stroke, peripheral arterial disease, coronary or peripheral revascularization (coronary bypass surgery, angioplasty of coronary arteries, or peripheral arterial disease-related angiography), or carotid endarterectomy or angioplasty. We additionally excluded those randomized to the intervention arm of the Dietary Modification trial (n = 2,648) to avoid post-randomization diet changes associated with the low-fat diet intervention, yielding a final sample of 16,835 women for the secondary prospective analyses (**Figure 1**).

**Figure 1.**
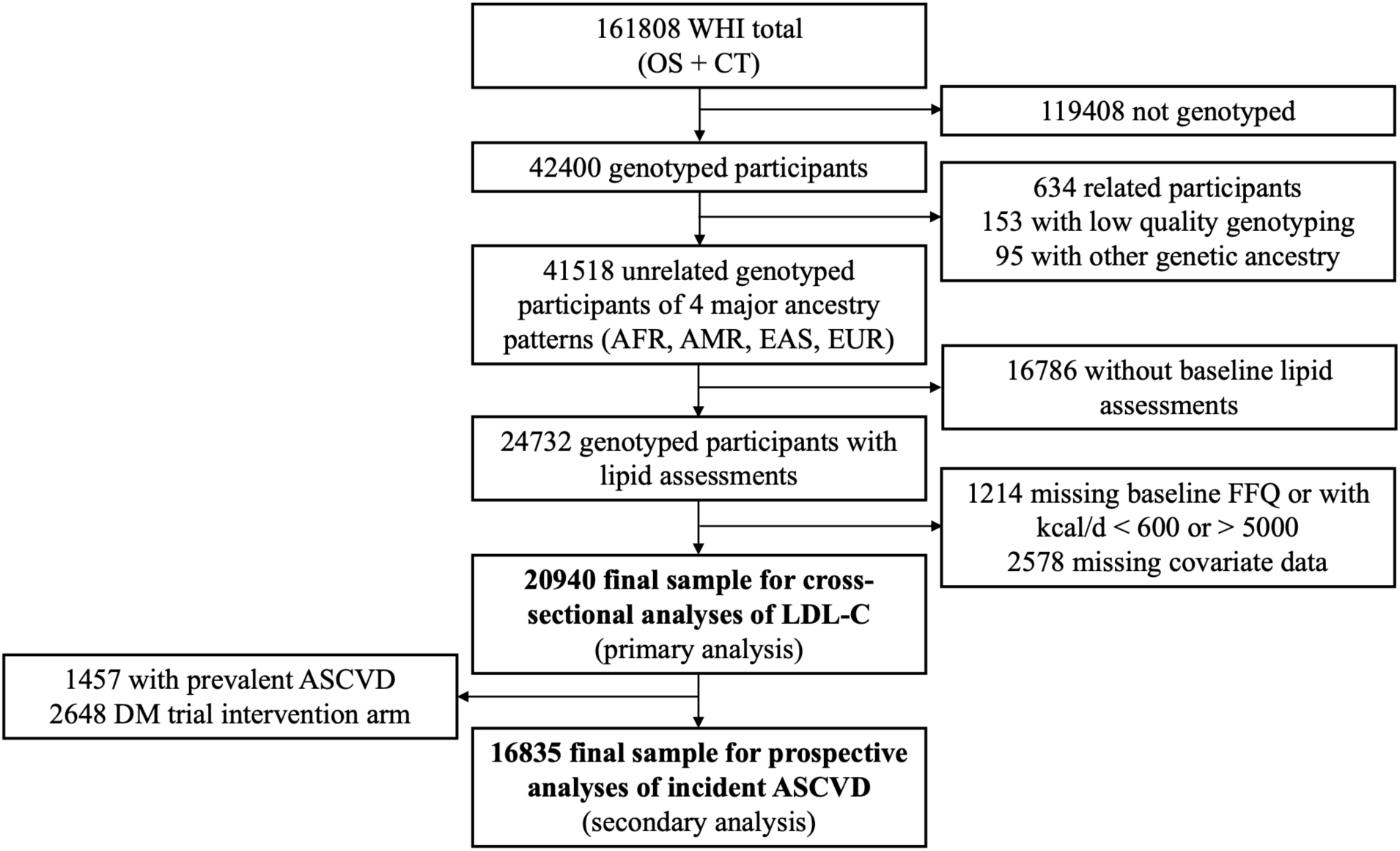
Study cohort flowchart. The study sample consisted of genotyped WHI participants who were unrelated and who had genetically inferred African (AFR), Admixed-American (AMR), East Asian (EAS), or European (EUR) ancestry and who had baseline measurement of LDL-C. Participants with missing or implausible FFQ data or missing covariates were excluded. For the prospective analysis of incident ASCVD, additional exclusions included participants with prevalent ASCVD and those randomized to the low-fat dietary intervention arm of the DM trial. ASCVD, atherosclerotic cardiovascular disease; CT, clinical trial; DM trial, Diet Modification trial. FFQ, food frequency questionnaire; LDL-C, low-density lipoprotein cholesterol; OS, observational study; WHI, Women’s Health Initiative.

### Genotyping data and polygenic score computation

Genotyping was performed using a range of arrays across the various ancillary studies, and all but two studies that used older array technology were included (**Supplementary Table 1**). All studies underwent imputation using the NHLBI Trans-Omics for Precision Medicine (TOPMed) imputation server, with TOPMed-r2 as the reference panel.^29^ Genetic ancestry was inferred within the *pgsc_calc* pipeline,^30^ using principal component analysis projection and assignment to ancestry groups based on genetic similarity to the 1000 Genomes Project Phase 3 super-populations.^31^ We used a previously validated multi-ancestry polygenic score for LDL-C (PGS000889) to quantify background genetic propensity towards higher or lower LDL-C, henceforth referred to as PGS_LDL_.^26^ The PGS_LDL_ was computed using *pgsc_calc*^30^ with a harmonized set of SNPs across all ancillary studies, resulting in 7,812 SNPs (86.7%) of the 9,009 autosomal variants included in the original PGS_LDL_.^26^ Lastly, the raw PGS_LDL_ was standardized in each ancestry group separately to have a mean of 0 and SD of 1. Accordingly, results are presented per 1-SD change in the score.

### Dietary assessment

Baseline dietary intake, collected concurrently with LDL-C measurements, was assessed via a semi-quantitative FFQ developed and validated for the WHI, which included additional items to enhance sensitivity for fat intake assessment.^32^ Participants were asked to report usual intake frequency and portion size for 122 foods and beverages, reflecting dietary consumption over the previous 3 months. Nutrient-level data were derived from the FFQ using the Nutrition Data System for Research software (version 2005; Nutrition Coordinating Center, University of Minnesota, Minneapolis, MN),^33^ with macronutrients (saturated fat, unsaturated fat, trans fat, protein, and carbohydrates) expressed as a percentage of total daily energy intake. In prior validation studies within this cohort, the FFQ demonstrated Pearson correlation coefficients of 0.63 for saturated fat and 0.67 for carbohydrates (modeled substitution variables, as described in the Statistical Analyses) when compared with the mean of eight records and recalls (four 24-hour recalls and one 4-day food record).^32^ Overall diet quality was assessed using the Alternative Healthy Eating Index (AHEI-2010), calculated following standard scoring procedures.^34^ Total scores range from 0 to 110, with higher scores reflecting greater adherence to a healthy dietary pattern.

### Primary and secondary outcomes

Baseline LDL-C (primary outcome) was calculated from fasting (≥ 12 hours) measurements of total cholesterol, high-density lipoprotein cholesterol, and triglycerides using the Friedewald equation (n = 20,292; 96.1%)^35^ or measured directly using a homogeneous assay (Roche Diagnostics; n = 648; 3.1%). For participants who reported using lipid lowering therapy (statins, bile acid sequestrants, niacin, or fibrates), untreated LDL-C was estimated by dividing the measured value by 0.7.^36–38^ The secondary outcome was incident ASCVD, which was broadly defined as the composite of myocardial infarction, coronary revascularization, peripheral artery disease, ischemic stroke, and definite coronary heart disease death. Because this composite outcome includes revascularization procedures which may have been elective, we also examined a stricter composite outcome of myocardial infarction, ischemic stroke, and definite coronary heart disease death. Outcome adjudication in the WHI has been previously described.^39^

### Covariates

Covariates assessed at enrollment included age, self-reported race and ethnicity, body mass index, education (categorized as having less than a college degree vs. having a college/advanced degree), smoking history (categorized as current vs. past or never smoker), recreational physical activity (metabolic-hour equivalents per week), following a low-fat or low-cholesterol diet (Yes vs. No, asked separately from the FFQ), history of diabetes, history of ASCVD, lipid lowering therapy use, and estrogen therapy use. Self-reported race and ethnicity were: non-Hispanic White, non-Hispanic Black or African American, and Hispanic. All remaining participants were pooled into “Other” due to limited sample sizes. Additional covariates include WHI study arm (CT vs. OS), WHI ancillary study to account for potential differences in genotyping array, and the first 5 genetic principal components (PCs) to control for population structure. Detailed descriptions and reliability of baseline assessments have been previously published.^40^

### Statistical analysis

Continuous variables are reported as the median [Q1, Q3] and categorical variables are expressed as percentages. Participants were stratified by polygenic score as having a PGS_LDL_ below or above the median for primary analyses unless otherwise specified. Differences in descriptive variables between PGS_LDL_ groups were assessed using the Kruskal-Wallis test for continuous variables or Chi-squared tests for categorical variables. PGS_LDL_ performance across ancestry groups was assessed by effect sizes (β coefficients) and model improvement (incremental R²). The 95% confidence intervals for the incremental R^2^ were derived using percentile bootstrapping. Multivariable linear regression models were used to evaluate the associations of PGS_LDL_ and saturated fat intake with LDL-C and to test their interaction. Reported *P*-values for interaction were obtained using likelihood ratio tests comparing nested models with and without the interaction term.

Because macronutrient intakes are interdependent under a stable total energy intake (i.e., an increase in one corresponds to a decrease in another), the association between saturated fat intake and LDL-C was estimated using leave-one-out isocaloric macronutrient substitution models.^41^ These models control for total energy intake and estimate the effect of replacing a specified proportion of energy from one macronutrient with another.^41^ For this study, an isocaloric substitution of 5% of total energy from carbohydrates with saturated fat was modeled, with saturated fat, unsaturated fat, trans fat, protein, and alcohol included in the model and leaving carbohydrates out to define the substitution.

For the secondary analyses of incident ASCVD, Cox proportional hazards models were used to estimate the associations of PGS_LDL_ and saturated fat intake with 10-year risk for incident ASCVD and to evaluate their multiplicative and additive interactions. The multiplicative interaction was assessed using likelihood ratio tests comparing nested models with and without the interaction term. The additive interaction was quantified using the relative excess risk due to interaction (RERI), with the 95% confidence intervals estimated using the delta method.^42^ Follow-up time for non-cases was defined as time from enrollment to last follow-up, 10 years, or death not due to coronary heart disease, whichever occurred first and was defined as time from enrollment to first ASCVD event for cases. The proportional hazards assumption was assessed using Schoenfeld residuals, with no evidence of violations.

Restricted cubic splines with three knots, implemented via the InteractionRCS package in R,^43^ were used to visualize the pattern of association between saturated fat and each outcome across the PGS_LDL_ spectrum. To generate these plots, the PGS_LDL_ was modeled using a restricted cubic spline with interaction terms between saturated fat intake and the spline basis functions. Separately, to formally test for nonlinear interaction, models were fit including a restricted cubic spline term for the product of saturated fat intake and PGS_LDL_. Because the primary objective of this analysis was to evaluate whether the interaction itself departed from linearity, we focused the statistical test on the nonlinear component of the interaction. *P*-values for nonlinear interaction were obtained using likelihood ratio tests comparing the full model to a reduced additive model without the spline interaction term. All models adjusted for covariates described above, including age, age^2^ (primary analyses only), body mass index, education, smoking status, physical activity, following a low-fat or low-cholesterol diet, history of diabetes, history of ASCVD (primary analyses only), lipid lowering therapy use, estrogen therapy use, self-reported race and ethnicity, WHI study type, genotyping array, and genetic PCs 1-5. Significance was defined at *P*<0.05. Analyses were performed using R version 4.3.3 (The R Foundation for Statistical Computing).

### Sensitivity analyses

For LDL-C, we conducted sensitivity analyses in the subset of participants without a history of diabetes, in those without a history of ASCVD, in those not using lipid-lowering therapy at baseline, and in those who did not report a low-fat or low-cholesterol diet at baseline. We also performed analyses stratified by genetically inferred ancestry groups. Further, because the multi-ancestry PGS_LDL_ used in this study (PGS000889)^26^ was derived from genome-wide association study (GWAS) that included a subset of ∼28K WHI participants (∼2.6% of the total discovery sample; n ∼ 1.09M), we replicated our analysis using an alternative polygenic score for LDL-C (PGS002654).^44^ This score was derived from a European-ancestry GWAS and that did not include WHI participants.

For analyses of incident ASCVD, all Cox models were re-evaluated after excluding participants using lipid-lowering therapy at baseline and in models incorporating lipid-lowering therapy modeled as a time-dependent covariate, with treatment status updated throughout follow-up to reflect initiation and discontinuation of therapy. Additional sensitivity analyses included models with an expanded set of covariates, including five-category education variable (less than high school, high school/GED or vocational, some college/associates degree, bachelor’s degree, graduate/ professional degree), six-category income variable (< $20,000, $20,000 - $34,999, $35,000 - $49,999, $50,000 - $74,999, ≥ 75,000, unknown or missing), and four-category smoking variable (never or past smoker, current smoker: < 5 pack years, current smoker: 5 to < 20 pack years, current smoker: ≥ 20 pack years). For analyses requiring categorization of saturated fat intake, which were not conducted within the isocaloric substitution framework, the expanded models additionally adjusted for overall diet quality using the AHEI.

## Results

### Study cohort

At baseline, the median age was 65.0 years [Q1: 58.0, Q3: 70.0], and 47.9% of individuals self-identified as non-Hispanic White, 33.0% as non-Hispanic Black, and 14.4% as Hispanic. The median LDL-C, corrected for lipid lowering therapy when present, was 148.0 mg/dL [123.0, 175.7], with 9% of participants on lipid-lowering therapy at baseline (**Table 1**). Compared to the full WHI cohort, our study subset had a greater representation of individuals who self-identified as Black or Hispanic (**Supplementary Table 2**). This enrichment aligns with the design of the WHI ancillary studies, which targeted genotyping for populations and outcomes of interest (**Supplementary Table 1**).

**Table 1.** Baseline characteristics of the study population.

|  | **All**<br>**(n = 20,940)** | **PGS<sub>LDL</sub> < median**<br>**(n = 10,469)** | **PGS<sub>LDL</sub> ≥ median**<br>**(n = 10,471)** | **P-**<br>**value** |
| --- | --- | --- | --- | --- |
| Untreated LDL-C, mg/dL | 148.0 [123.0, 175.7] | 135.0 [114.0, 160.0] | 162.0 [137.0, 188.2] | <2e-16 |
| Age, years | 65.0 [58.0, 70.0] | 65.0 [58.0, 70.0] | 65.0 [58.0, 70.0] | 0.70 |
| Self-reported race and ethnicity [n (%)] |  |  |  | 0.18 |
| Non-Hispanic White | 10027 (47.9) | 4989 (47.7) | 5038 (48.1) |  |
| Non-Hispanic Black | 6910 (33.0) | 3522 (33.6) | 3388 (32.4) |  |
| Hispanic | 3008 (14.4) | 1467 (14.0) | 1541 (14.7) |  |
| Other | 995 (4.8) | 491 (4.7) | 504 (4.8) |  |
| Genetically inferred ancestry [n (%)] |  |  |  | 1 |
| European (EUR) | 10488 (50.1) | 5244 (50.1) | 5244 (50.1) |  |
| African (AFR) | 7046 (33.6) | 3522 (33.6) | 3524 (33.6) |  |
| Admixed American (AMR) | 2996 (14.3) | 1498 (14.3) | 1498 (14.3) |  |
| East Asian (EAS) | 410 (2.0) | 205 (2.0) | 205 (2.0) |  |
| College degree or higher [n (%)] | 6994 (33.4) | 3601 (34.4) | 3393 (32.4) | 0.002 |
| History of ASCVD [n (%)] | 1457 (7.0) | 688 (6.6) | 769 (7.3) | 0.03 |
| History of diabetes [n (%)] | 1941 (9.3) | 1016 (9.7) | 925 (8.8) | 0.03 |
| Lipid lowering drug use [n (%)] | 1891 (9.0) | 600 (5.7) | 1291 (12.3) | <2e-16 |
| Estrogen use [n (%)] | 2679 (12.8) | 1355 (12.9) | 1324 (12.6) | 0.53 |
| BMI, kg/m<sup>2</sup> | 28.6 [25.1, 32.9] | 28.6 [25.1, 32.9] | 28.5 [25.0, 32.8] | 0.15 |
| Obesity (≥ 30.0 kg/m<sup>2</sup>) | 8457 (40.4) | 4261 (40.7) | 4196 (40.1) | 0.36 |
| Current smoker [n (%)] | 1887 (9.0) | 972 (9.3) | 915 (8.7) | 0.18 |
| Physical activity, MET-h/week | 6.4 [1.3, 15.6] | 6.5 [1.3, 15.5] | 6.4 [1.5, 15.8] | 0.26 |
| Following a low-fat diet [n (%)] | 7614 (36.4) | 3353 (32.0) | 4261 (40.7) | <2e-16 |
| Clinical trial participation [n (%)] | 15,570 (74.4) | 7869 (75.2) | 7701 (73.5) | 0.008 |
| AHEI-2010 | 49.3 [42.6, 56.5] | 49.2 [42.4, 56.5] | 49.4 [42.7, 56.5] | 0.07 |

Values are reported as the median [Q1, Q3] or n (%). *P*-values compare genetic risk groups (PGS<sub>LDL</sub> < vs. ≥ median) and are from Kruskal-Wallis test for continuous variables or Chi-squared tests for categorical variables. AHEI-2010, Alternative Healthy Eating Index; ASCVD, atherosclerotic cardiovascular disease; BMI, body mass index; LDL-C, low-density lipoprotein cholesterol; MET-h/week, metabolic equivalent hours per week. PGS<sub>LDL</sub>, polygenic score for low-density lipoprotein cholesterol.

Descriptive characteristics of the study population stratified by PGS_LDL_ below or above the population median were also evaluated (**Table 1**). Participants with a high PGS_LDL_ (PGS_LDL_ ≥ median) had significantly higher baseline LDL-C levels, a greater prevalence of ASCVD, and a lower prevalence of diabetes,^45^ compared to those with a low PGS_LDL_ (PGS_LDL_ < median).

Additionally, a greater proportion of participants with a high PGS_LDL_ reported being on lipid lowering therapy and following a low-fat or low-cholesterol diet at baseline. No differences in diet quality, assessed by the AHEI, were observed between genetic risk groups. The PGS_LDL_ was effectively normalized across ancestry groups (**Supplementary Figure 1**), and the normalized score was not predictive of genetically inferred ancestry or self-reported race and ethnicity (**Table 1**).

### Individual and combined associations of polygenic score and saturated fat with LDL-C

The association between the normalized PGS_LDL_ and measured LDL-C was assessed in the pooled cohort (**Table 2**) and separately by ancestry groups (**Supplementary Table 3**). The PGS_LDL_ was strongly associated with LDL-C, with a 14.9 mg/dL increase in LDL-C per 1-SD increase in the PGS_LDL_ (**Table 2**). In terms of both effect size (beta) and model improvement (incremental R^2^), similar performance in the EUR and AFR groups, a modest attenuation in the AMR group, and a considerable attenuation in the EAS group were observed (**Supplementary Table 3**), consistent with previous assessments of this polygenic score.^26^ The distribution of saturated fat intake and correlations between macronutrients (expressed as % of total daily calories), AHEI, and PGS_LDL_ in the pooled cohort are shown in **Supplementary Figure 2**. The median % of total calories from saturated fat in the pooled cohort was 10.9% [Q1: 8.8, Q3: 13.0]. Saturated fat was significantly associated with LDL-C, such that a 5% increase in calories from saturated fat, in place of carbohydrates, was associated with a 1.73 mg/dL higher LDL-C (**Table 2**), an effect size consistent with a prior cross-sectional substitution analysis.^46^ The associations between the PGS_LDL_ and saturated fat intake with LDL-C were independent of one another in a combined (additive) model that included both exposures (**Table 2**). In fact, with adjustment for PGS_LDL_, the association between saturated fat and LDL-C strengthened. There was minimal correlation between saturated fat intake and PGS_LDL_ (r = -0.06; **Supplementary Figure 2**).

**Table 2.** Individual and combined associations of saturated fat intake and polygenic score with baseline LDL-C.

|  | **Independent variable** | **$\beta$ (SE)<sup>1</sup>** | **P-value** |
| --- | --- | --- | --- |
| **Individual models** | Saturated fat | 1.73 (0.57) | 0.003 |
|  | PGS<sub>LDL</sub> | 14.90 (0.25) | <2e-16 |
| **Combined model** | Saturated fat | 2.32 (0.53) | 1.2e-5 |
|  | PGS<sub>LDL</sub> | 14.92 (0.25) | <2e-16 |

Models adjust for age, age<sup>2</sup>, genetic principal components 1-5, WHI ancillary study, body mass index, self-reported race and ethnicity, education, smoking, metabolic hour equivalents per week, history of atherosclerotic cardiovascular disease, history of diabetes, lipid-lowering therapy, estrogen therapy, and following a low-fat diet. Individual models evaluate saturated fat and PGS<sub>LDL</sub> separately; The combined model evaluates saturated fat and PGS<sub>LDL</sub> together in an additive model.
<sup>1</sup>The $\beta$ (SE) for saturated fat represents the change in LDL-C (mg/dL) change per 5% of total calories increase in saturated fat in place of carbohydrates. The $\beta$ (SE) for the PGS<sub>LDL</sub> represents the change in LDL-C (mg/dL) per 1-SD unit increase in the PGS<sub>LDL</sub>.

### Interaction between polygenic score and saturated fat on LDL-C

We stratified the cohort by PGS_LDL_ above or below the median and examined the association between saturated fat and LDL-C within each stratum. We observed a difference in association by PGS_LDL_ strata (*P*-interaction = 0.01), with an attenuation in the association among those with PGS_LDL_ below the median (**Figure 2A**). We used spline analysis to visualize how the association between saturated fat intake and LDL-C varied across the full polygenic risk spectrum (**Figure 2B**). At the lower end of the PGS_LDL_ distribution, the association between saturated fat and LDL-C remained modest and relatively stable, but at higher PGS_LDL_, the effect size showed an upward slope. Consistent with this pattern, a separate formal test of nonlinear interaction showed that allowing the interaction between saturated fat intake and PGS_LDL_ to be nonlinear significantly improved model fit (*P*-interaction, nonlinear = 0.01).

**Figure 2.**
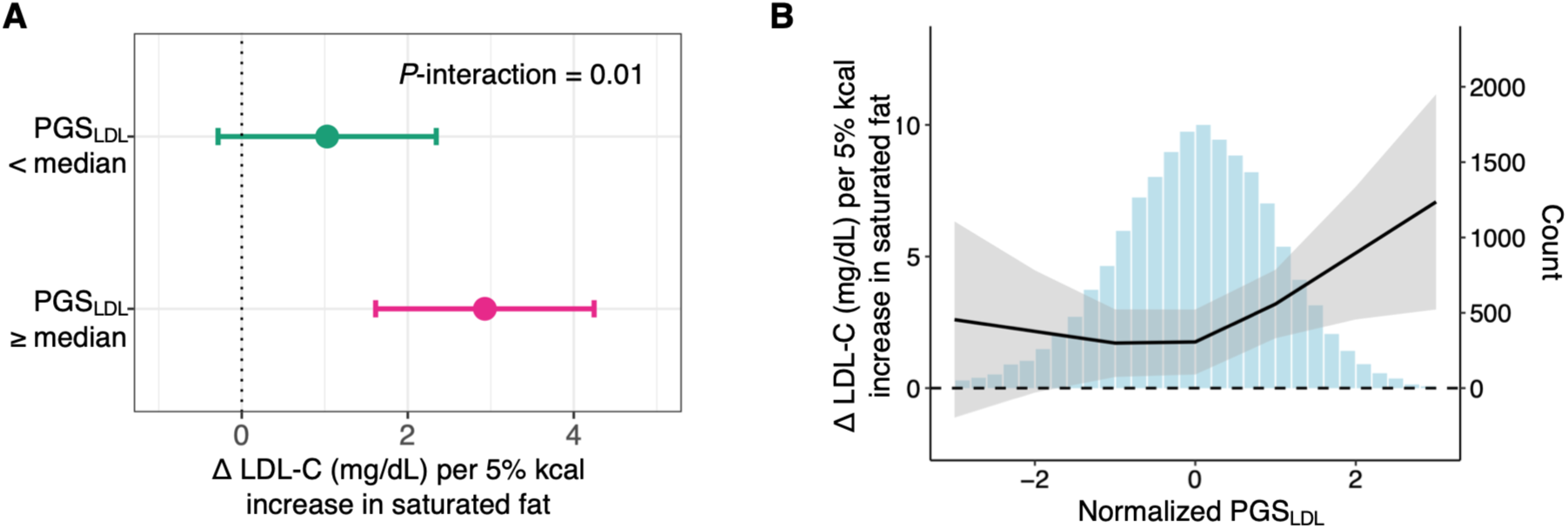
Polygenic score modifies the association between saturated fat intake and low-density lipoprotein cholesterol. (A) Forest plot showing the β (95%CI) representing the estimated difference in LDL-C (mg/dL) associated with each 5% of total energy increase in saturated fat in place of carbohydrates, stratified by PGS_LDL_ category. (B) Restricted cubic spline showing the estimated change in LDL-C associated with each 5% of total energy from saturated fat replacing carbohydrates (left y-axis) across the genetic risk spectrum (x-axis). The histogram is referenced to the right y-axis and depicts the distribution of participants across the PGS_LDL_ spectrum. All models adjust for age, age^2^, genetic principal components 1-5, ancillary study, body mass index, self-reported race and ethnicity, education, smoking, metabolic hour equivalents per week, history of atherosclerotic cardiovascular disease, history of diabetes, lipid-lowering therapy, estrogen therapy, and following a low-fat diet. LDL-C, low-density lipoprotein cholesterol; PGS_LDL_, polygenic score for LDL-C.

### Sensitivity analyses for the interaction between polygenic score and saturated fat on LDL-C

Given the evidence of a nonlinear interaction, we additionally examined quartiles of PGS_LDL_ to assess whether finer stratification better characterized the interaction (**Supplementary Figure 3**). Associations between saturated fat intake and LDL-C were similar within the lower half of the PGS_LDL_ distribution (Q1 vs Q2, P = 0.29) and within the upper half (Q3 vs Q4, P = 0.82), indicating that further stratification did not provide additional insight beyond the median-based approach.

We next repeated our analyses in several subsets of participants, including those without a history of diabetes, those without a history of ASCVD, those not on lipid-lowering therapy, and those not following a low-fat or low-cholesterol diet at baseline. For all analyses, we observed the same general pattern of an attenuated association within the low PGS_LDL_ strata (**Figure 3**).

**Figure 3.**
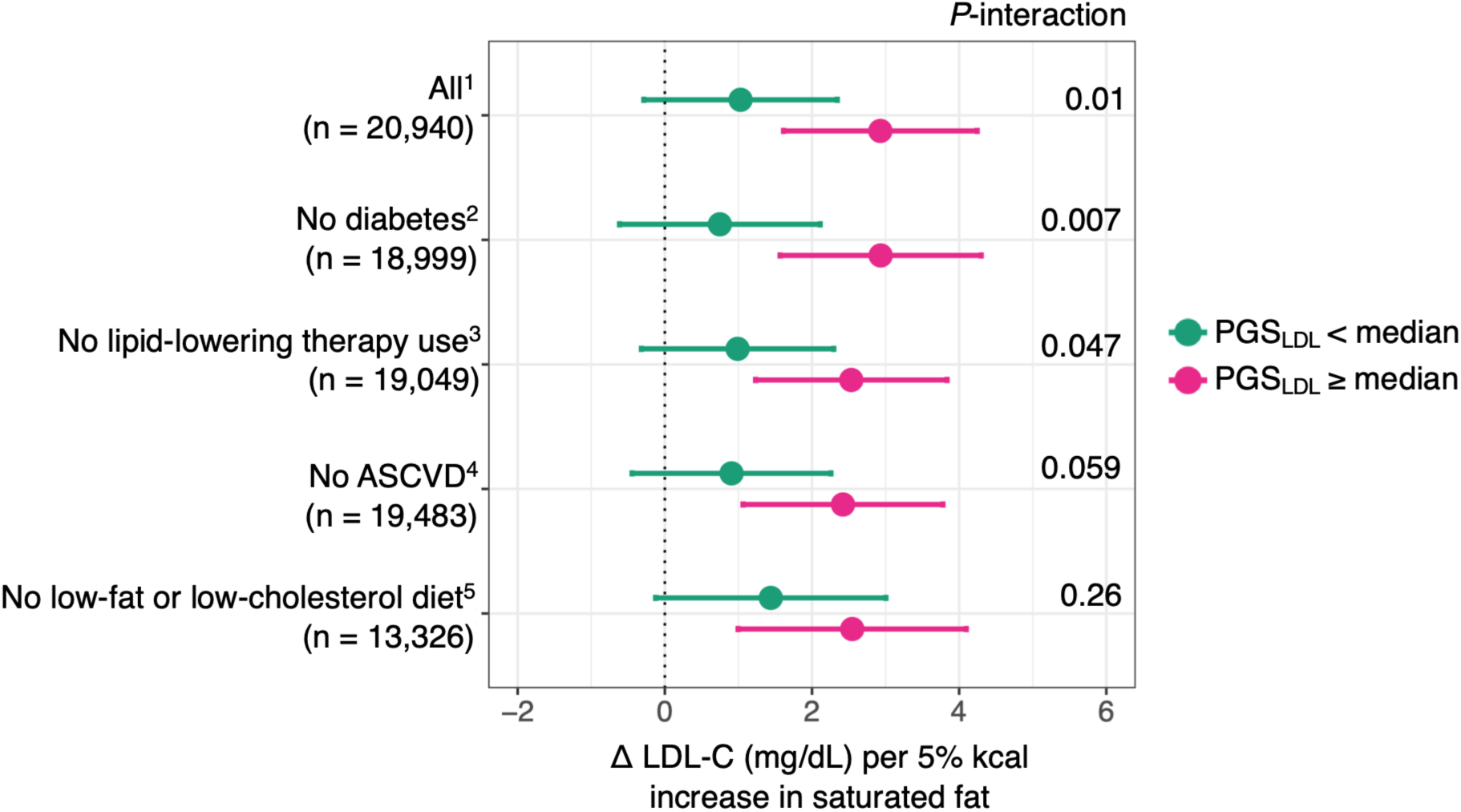
Sensitivity analyses of the interaction between polygenic score and saturated fat intake on low-density lipoprotein cholesterol. Forest plot showing the β (95% CI) representing the estimated change in LDL-C (mg/dL) associated with each 5% of total energy increase in saturated fat in place of carbohydrates, stratified by PGS_LDL_ category. All models adjust for age, age^2^, genetic principal components 1-5, ancillary study, body mass index, self-reported race and ethnicity, education, smoking, metabolic hour equivalents per week, and estrogen therapy. ASCVD, atherosclerotic cardiovascular disease; PGS_LDL_, polygenic score for low-density lipoprotein cholesterol. ^1^Models additionally adjust for history of diabetes, history of ASCVD, lipid lowering therapy, and following a low-fat diet. Low PGS_LDL_: n = 10,469; High PGS_LDL_: n = 10,471. ^2^Models additionally adjust for history of ASCVD, lipid lowering therapy, and following a low-fat diet. Low PGS_LDL_: n = 9,498; High PGS_LDL_: n = 9,501. ^3^Models additionally adjust for history of diabetes, history of ASCVD, and following a low-fat diet. Low PGS_LDL_: n = 9,522; High PGS_LDL_: n = 9,527. ^4^Models additionally adjust for history of diabetes, lipid lowering therapy, and following a low-fat diet. Low PGS_LDL_: n = 9,741; High PGS_LDL_: n = 9,742. ^5^Models additionally adjust for history of diabetes, history of ASCVD, and lipid lowering therapy. Low PGS_LDL_: n = 6,633; High PGS_LDL_: n = 6,633.

We further examined the robustness of the interaction to ancestry-specific differences. In the pooled cohort, inclusion of ancestry-by-saturated fat intake and ancestry-by-PGS_LDL_ interaction terms had minimal impact on the interaction estimate (β = 1.69 mg/dL; 95% CI, 0.16 to 3.22) compared with the primary model without these terms (β = 1.90 mg/dL; 95% CI, 0.37 to 3.43). Likewise, a random-effects meta-analysis of the ancestry-specific interaction estimates yielded a comparable pooled estimate (β = 1.68 mg/dL; 95% CI, 0.16 to 3.19), with no evidence of between-ancestry heterogeneity (**Supplementary Figure 4**). Together, these findings indicate that the observed interaction was robust to alternative approaches accounting for ancestry-specific effects. However, although the ancestry-stratified analyses were limited by small sample sizes, the patterns observed suggest that the interaction may be predominantly driven by the two populations for which the PGS_LDL_ showed best performance (EUR and AFR; **Supplementary Figure 4**).

We then evaluated whether the interaction pattern we observed in this study is specific to the polygenic score by repeating the analysis using an alternative score (PGS002654), which was derived from a European-ancestry GWAS. In the EUR subset, both our primary polygenic score and the alternative polygenic score showed a similar pattern to what we observed in our main analyses: a relatively stable and modest association at the lower end of the genetic spectrum but upward-sloping effect sizes at the higher end (**Supplementary Figure 5**).

Lastly, to determine if the observed interaction is specific to saturated fat, we performed our analysis using the other macronutrients in the substitution model (**Supplementary Figure 6**).

None showed a significant interaction, though the interaction with trans fat was borderline (*P*-interaction = 0.05).

### The associations of polygenic score and saturated fat with incident ASCVD

Among 16,835 participants without prevalent ASCVD at baseline, 1,582 (9.4%) experienced incident ASCVD within 10 years (median follow-up: 10 years). Baseline characteristics of participants included in the incident ASCVD analyses are shown in **Supplementary Table 4**.

Unlike the LDL-C analyses, in which the median-based stratification adequately characterized the interaction, quartile analysis for ASCVD demonstrated that the association between saturated fat intake and ASCVD risk was largely confined to individuals in the highest PGS_LDL_ quartile (Q4), whereas similar attenuated associations were observed across quartiles Q1-Q3 (**Supplementary Figure 7**). Based on this pattern, subsequent analyses compared participants in PGS_LDL_ Q4 with those in Q1-Q3. In fully adjusted models, saturated fat intake was associated with a higher risk of incident ASCVD among participants in Q4 of PGS_LDL_ but not among those in Q1-Q3, demonstrating evidence of a multiplicative interaction (*P*-interaction = 0.006; **Figure 4A**). Similar findings were observed when evaluating the stricter composite outcome of myocardial infarction, ischemic stroke and coronary heart disease death (*P*-interaction = 0.01; **Figure 4B**).

**Figure 4.**
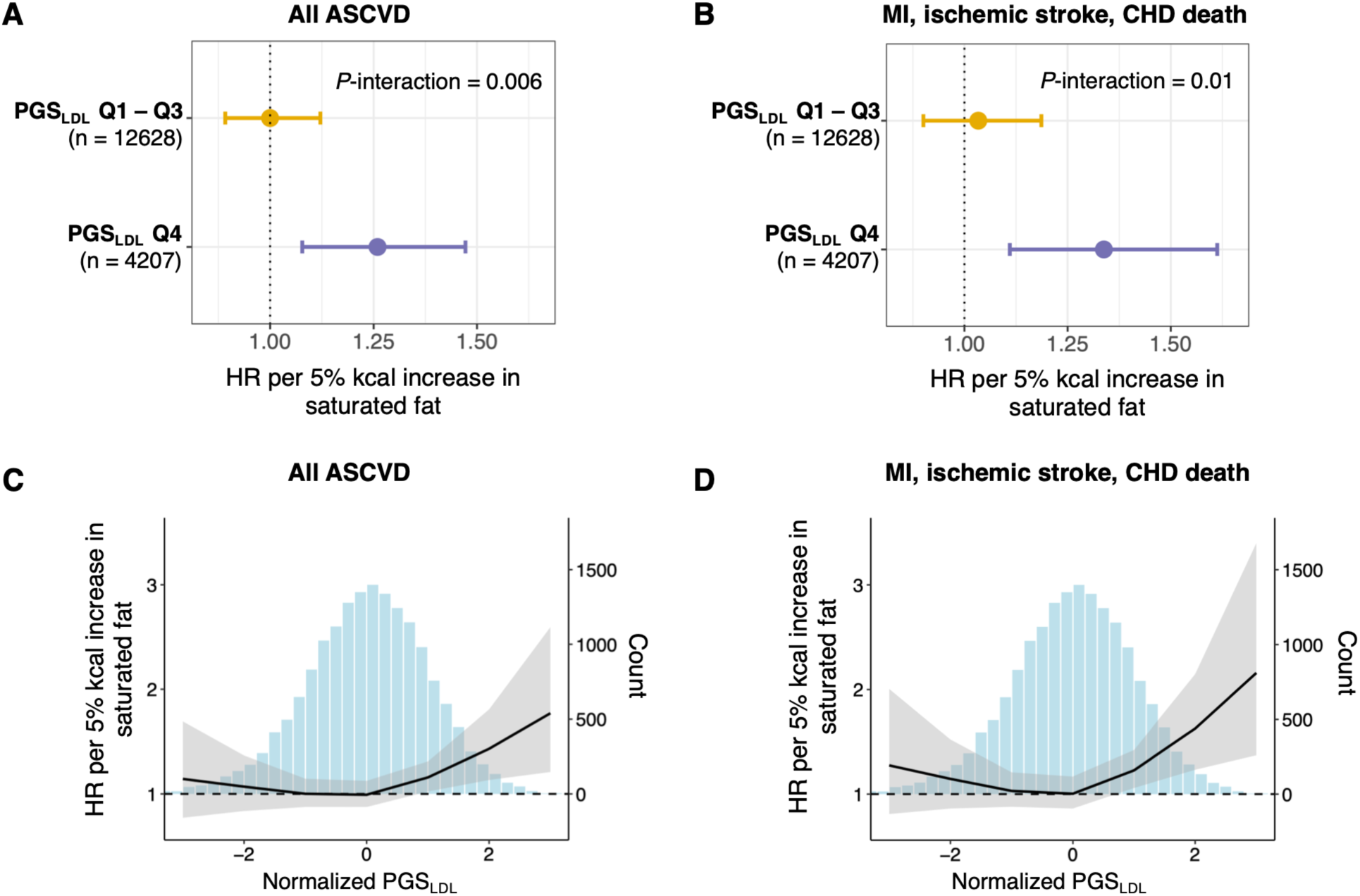
The association of saturated fat intake with incident ASCVD according to polygenic score. Forest plots show the hazard ratio (HR) and 95% CI for (A) all ASCVD and (B) the strict composite of myocardial infarction, ischemic stroke, and coronary heart disease death, associated with each 5% of total energy increase in saturated fat in place of carbohydrates by PGS_LDL_ category; Q4 indicates the highest quartile of PGS_LDL_, and Q1-Q3 indicate the lower three quartiles. Restricted cubic spline plots show the estimated HR and 95% CI for (C) all ASCVD or (D) the strict composite of myocardial infarction, ischemic stroke, and coronary heart disease death, associated with each 5% of total energy increase in saturated fat in place of carbohydrates (left y-axis) across the PGS_LDL_ spectrum (x-axis). The histogram in panels C and D, referenced to the right y-axis, depicts the distribution of participants across the PGS_LDL_ spectrum. All analyses are adjusted for age, genetic principal components 1-5, ancillary study, body mass index, self-reported race and ethnicity, education, smoking, metabolic hour equivalents per week, history of diabetes, lipid lowering therapy, estrogen therapy, and following a low-fat diet. ASCVD, atherosclerotic cardiovascular disease; PGS_LDL_, polygenic score for low-density lipoprotein cholesterol.

We next used spline analysis to visualize how the association between saturated fat intake and risk for incident ASCVD varied across the full spectrum of polygenic risk. For both the broad ASCVD outcome and the strict composite of myocardial infarction, ischemic stroke, and coronary heart disease death, the spline plots were consistent with the quartile-based analyses, showing relatively stable estimated effects across the lower end of the PGS_LDL_ distribution with larger estimated effects only observed at the higher end (**Figure 4**). We then performed separate formal tests of nonlinear interaction. For the broad definition of ASCVD, we did not find statistical evidence that allowing the interaction to be nonlinear improved model fit (*P*-interaction, nonlinear = 0.18). In contrast, evidence of a nonlinear interaction was observed for the strict endpoint (*P*-interaction, nonlinear = 0.03). Findings were consistent in analyses restricted to participants not using lipid-lowering therapy at baseline (**Supplementary Figure 8**), analyses incorporating lipid-lowering therapy as a time-varying covariate (**Supplementary Figure 9**), and analyses with additional adjustment for an expanded set of covariates (**Supplementary Figure 10**).

To further illustrate the joint associations of polygenic risk and saturated fat intake with incident ASCVD risk, we stratified the population by both PGS_LDL_ (high = Q4 vs. low = Q1-Q3) and saturated fat intake (% calories from saturated fat; high = Q4 vs. low = Q1-Q3), creating a 2-by-2 cross-classification. Descriptive cumulative incidence plots demonstrated a stepwise increase in the 10-year incidence of ASCVD (**Figure 5A**). The 10-year incidence was 9.5% [95% CI: 8.9, 10.1] for individuals with low PGS_LDL_ and low saturated fat intake, 10.4% [95% CI: 9.5, 11.5] for those with low PGS_LDL_ and high saturated fat intake, 9.8% [95% CI: 8.7, 10.8] for those with high PGS_LDL_ and low saturated fat intake, and 14.3% [95% CI: 12.1, 16.7] for individuals with both high PGS_LDL_ and high saturated fat intake. A similar gradient was observed when restricting to the composite of myocardial infarction, ischemic stroke, and coronary heart disease death, with corresponding incidences of 6.5% [95% CI: 6.0, 7.0], 7.2% [95% CI: 6.3, 8.1], 6.4% [95% CI: 5.6, 7.3], and 9.9% [95% CI: 8.1, 12.0], respectively (**Figure 5B**). Cumulative incidence plots accounting for non-coronary heart disease deaths as competing risks are shown in **Supplementary Figure 11**.

**Figure 5.**
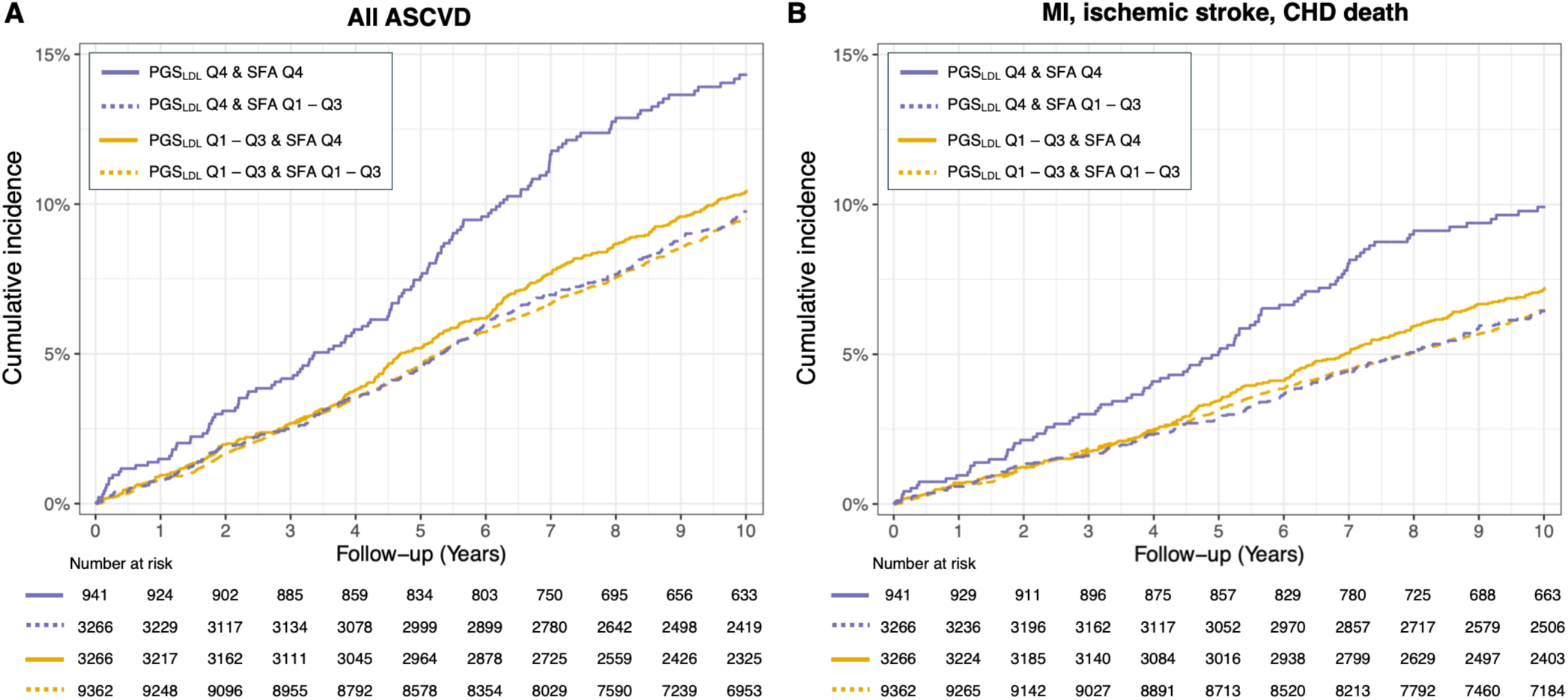
Cumulative incidence curves (1 – Kaplan-Meier estimates) for (A) all ASCVD and (B) the strict composite of myocardial infarction, ischemic stroke, and coronary heart disease death, according to joint categories of polygenic score and saturated fat intake. For both PGS_LDL_ and saturated fat intake, Q4 denotes the highest quartile, and Q1-Q3 indicate the lower three quartiles. Corresponding cumulative incidence curves accounting for competing risks are shown in Supplementary Figure 11. ASCVD, atherosclerotic cardiovascular disease; PGS_LDL_, polygenic score for low-density lipoprotein cholesterol; SFA, saturated fat intake (as % of total daily energy intake).

We then quantified these joints associations using adjusted Cox models and evaluated additive interactions using RERI. In a fully adjusted model using participants with both low PGS_LDL_ and low saturated fat intake as the reference, those with both high PGS_LDL_ and high saturated fat intake had a 45% higher risk of incident ASCVD (**Table 3**; Model 1). This association remained significant after further adjustment for baseline untreated LDL-C (**Table 3**; Model 2A) or measured LDL-C (**Table 3**; Model 2B). Similar effect estimates were observed for the strict composite of myocardial infarction, ischemic stroke, and coronary heart disease death (**Table 3**). Evidence of additive interaction was observed for both the broad (RERI = 0.37 [95% CI, 0.06 to 0.67]) and strict (RERI = 0.41 [95% CI, 0.04 to 0.78]) ASCVD outcomes (**Table 3**). Associations remained largely unchanged in analyses restricted to participants not using lipid-lowering therapy at baseline (**Supplementary Table 5**), analyses incorporating lipid-lowering therapy as a time-varying covariate (**Supplementary Table 5**), and analyses with additional adjustment for an expanded set of covariates (**Supplementary Table 6**). Likewise, findings remained consistent when accounting for non-coronary heart disease deaths as competing risks using Fine-Gray models (**Supplementary Table 7**). Finally, to evaluate the robustness of the findings to the method used to categorize saturated fat intake, we repeated the analyses using a median split rather than the upper quartile, which yielded consistent although slightly attenuated results (**Supplementary Table 8**).

**Table 3.** Joint associations of polygenic score and saturated fat intake on incident ASCVD.

| All ASCVD |  |  |  |  |  |  |  |  |
| --- | --- | --- | --- | --- | --- | --- | --- | --- |
| Group | N |  | Model 1 |  | Model 2A |  | Model 2B |  |
|  | Total | Cases | HR [95% CI] | P | HR [95% CI] | P | HR [95% CI] | P |
| PGS<sub>LDL</sub> Q1-Q3 & SFA Q1-Q3 | 8525 | 837 | Reference | - | Reference | - | Reference | - |
| PGS<sub>LDL</sub> Q1-Q3 & SFA Q4 | 2947 | 319 | 1.04 [0.91, 1.19] | 0.60 | 1.02 [0.90, 1.17] | 0.73 | 1.03 [0.90, 1.17] | 0.72 |
| PGS<sub>LDL</sub> Q4 & SFA Q1-Q3 | 2967 | 299 | 1.04 [0.91, 1.19] | 0.54 | 0.94 [0.82, 1.08] | 0.40 | 0.93 [0.81, 1.07] | 0.32 |
| PGS<sub>LDL</sub> Q4 & SFA Q4 | 814 | 127 | 1.45 [1.20, 1.75] | 0.0001 | 1.29 [1.06, 1.56] | 0.01 | 1.28 [1.05, 1.55] | 0.01 |
| Additive interaction (RERI) [95% CI] |  |  | 0.37 [0.06, 0.67] |  | 0.32 [0.04, 0.56] |  | 0.32 [0.04, 0.59] |  |
| Myocardial infarction, ischemic stroke, and coronary heart disease death |  |  |  |  |  |  |  |  |
| Group | N |  | Model 1 |  | Model 2A |  | Model 2B |  |
|  | Total | Cases | HR [95% CI] | P | HR [95% CI] | P | HR [95% CI] | P |
| PGS<sub>LDL</sub> Q1-Q3 & SFA Q1-Q3 | 8794 | 568 | Reference | - | Reference | - | Reference | - |
| PGS<sub>LDL</sub> Q1-Q3 & SFA Q4 | 3047 | 219 | 1.03 [0.88, 1.21] | 0.73 | 1.02 [0.87, 1.20] | 0.79 | 1.02 [0.87, 1.20] | 0.79 |
| PGS<sub>LDL</sub> Q4 & SFA Q1-Q3 | 3069 | 197 | 1.03 [0.87, 1.21] | 0.79 | 0.96 [0.81, 1.14] | 0.63 | 0.95 [0.80, 1.13] | 0.57 |
| PGS<sub>LDL</sub> Q4 & SFA Q4 | 853 | 88 | 1.47 [1.17, 1.84] | 0.001 | 1.37 [1.08, 1.72] | 0.009 | 1.36 [1.08, 1.72] | 0.01 |
| Additive interaction (RERI) [95% CI] |  |  | 0.41 [0.04, 0.78] |  | 0.38 [0.04, 0.73] |  | 0.38 [0.04, 0.73] |  |

Hazard ratio (HR) and 95% CI from Cox proportional hazards models of incident ASCVD. The RERI was calculated from the estimated HRs, with the 95% CIs estimated using the delta method. Model 1 is adjusted for age, genetic principal components 1-5, ancillary study, body mass index, self-reported race and ethnicity, education, smoking, metabolic hour equivalents per week, lipid-lowering therapy, estrogen therapy, and following a low-fat diet. Model 2 is a mediator-adjusted model: Model 2A adjusts for Model 1 covariates plus baseline untreated LDL-C (adjusted for lipid lowering therapy when present) and Model 2B adjusts for model 1 covariates plus baseline measured LDL-C (unadjusted for lipid lowering therapy). For both PGS<sub>LDL</sub> and saturated fat intake, Q4 denotes the highest quartile, and Q1-Q3 indicate the lower three quartiles. ASCVD, atherosclerotic cardiovascular disease; LDL-C, low density lipoprotein cholesterol; PGS<sub>LDL</sub>, polygenic score for low-density lipoprotein cholesterol; RERI, relative excess risk due to interaction; SFA, saturated fat intake (as % of total daily calories).

## Discussion

In this study, we found evidence that genetic propensity, as captured by a genome-wide polygenic score for LDL-C, modified the association between saturated fat intake and LDL-C. Strikingly, our analysis suggests that this effect modification was not a simple multiplicative interaction. Rather, the modifying effect of polygenic score appeared to emerge only at one end of the genetic spectrum. At the low end, the association between saturated fat and LDL-C was modest but consistent. At the high end, the magnitude of association rose steeply with increasing polygenic score. Our secondary analyses of ASCVD endpoints showed a similar overall pattern, raising the possibility that the interaction observed for LDL-C may also extend to clinical outcomes. These findings carry several important implications when placed into the context of prior observations.

A large body of literature supports the presence of gene-diet interactions influencing LDL-C. Genetic variants at several lipid-related loci have been implicated, including *APOE*, *PPARG*, *APOA5*, *TCF7L2*, and *LIPC*.^16–23^ While genetic variants in these loci provide important avenues for mechanistic investigation, the clinical importance of specific common variants is likely to remain limited because the proportion of the population that carries any given variant is low and the impact of any single variant is small. For this reason, the use of polygenic scores to capture gene-diet interactions is appealing. One early effort used WHI data to conduct a genome-wide interaction study (GWIS) to identify genetic variants that interact with the association of total fat intake and LDL-C.^25^ Using this GWIS, the authors constructed a diet-response polygenic score consisting of 1,760 common variants, and they showed that this score could predict response to a low-fat diet. This study was limited by its reliance on a small and under-powered GWIS (n = 7,050) and by a study design that combined the effects of both saturated and unsaturated fats. Nonetheless, it provided an important proof of concept that gene-diet interactions affecting LDL-C may extend far beyond a handful of variants. A more recent analysis using data from the UK Biobank found evidence of an interaction between a polygenic score and a low-carbohydrate, high-fat (LCHF) diet. The LCHF diet was defined as <25% energy from carbohydrates and >45% from total fat, and only ∼1% of UK Biobank participants with diet data exhibited this dietary pattern.^24^ The polygenic score consisted of 223 common variants associated with LDL-C. Among participants with a high polygenic score (top 20%), they found that the LCHF diet was associated with higher LDL-C compared with a non-LCHF dietary pattern, but no differences between diet groups were observed among the lower 80% of polygenic risk. This study provided further evidence that genetic propensity may interact with diet, but it focused on a somewhat extreme dietary pattern, and the analysis was limited to a single categorical comparison.

Our study addresses some of the key limitations of prior work. First, our analysis disentangles saturated fat from total fat. This distinction is important given the well-established opposing effects of unsaturated fat compared to saturated fat.^2,3^ The clinical importance of this distinction is reflected in guidelines, which consistently recommend reducing saturated fat intake and replacing it with unsaturated fats to lower LDL-C.^4–6,47^ Accordingly, in this study we focused specifically on saturated fat intake, while also accounting for the compositional nature of dietary data. Second, our study is substantially larger than prior studies and encompasses a diversity of dietary patterns typical for the U.S. population. Third, our PGS_LDL_ represents a more powerful genetic instrument than used in prior studies, having been derived from a multi-ancestry GWAS meta-analysis of more than 1 million individuals across the globe.^26^

Our study also provides distinctly new insights. We have shown for the first time that the shape of the interaction between saturated fat and genetic background may be non-linear, and our secondary analyses further suggest that this pattern may further extend to risk for ASCVD. The shape of this interaction carries two important clinical messages. First, although the magnitude of association between saturated fat and LDL-C diminished at lower PGS_LDL_, it remained positive throughout. Likewise, for ASCVD endpoints, increased saturated fat consumption never appeared to be protective. Thus, while the response to saturated fat may vary from person-to-person, our analyses suggest that this heterogeneity reflects varying degrees of detriment, with no evidence for a protective effect for any genetic subgroup. The second message is that a large portion of the population carrying LDL-C-raising variants had a stronger association between saturated fat intake and LDL-C. One possible interpretation is that these individuals may be more sensitive to saturated fat in their diet, contradicting the common perspective that, “diet doesn’t matter,” for people predisposed to hypercholesterolemia. Our study raises the possibility that diet is even more important for such patients. Together, these findings reinforce current dietary recommendations to limit saturated fat intake while providing a potential explanation for the well-recognized heterogeneity in LDL-C responses. Nonetheless, the clinical utility of using genetic information to guide dietary recommendations remains to be established.

Several limitations warrant consideration. First, the observational design precludes causal inference, and residual confounding may persist despite careful multivariable adjustment. Furthermore, our analysis of LDL-C relies on cross-sectional data. In this setting, the magnitude of association between saturated fat and LDL-C is expected to be weaker than what is observed in diet intervention trials, due to variability of free-living dietary exposures and measurement error in population-level dietary assessment. Our effect estimates were consistent with prior studies of similar design.^46,48^ Second, dietary intake was assessed at a single time point using self-report, introducing potential measurement error and limiting the ability to capture longitudinal changes in dietary habits. Such measurement error would be expected to attenuate associations and reduce power to detect gene-diet associations. Future studies in large, diverse genotyped cohorts with repeated dietary assessments and adjudicated cardiovascular outcomes are needed to address this important gap and build on these findings. Third, the study population was restricted to postmenopausal women, which may limit the generalizability of these findings to other populations. Because both the effects of genetic variants on LDL-C ^49^ and the lipid responses to diet^50,51^ may differ by sex, future studies are needed to determine whether the observed gene-diet interaction is consistent across sexes and age groups. Fourth, we used the current best available polygenic score for LDL-C, with strong comparable validity in the two largest ancestry groups represented in our cohort (AFR and EUR). However, performance characteristics of this score are weaker for the AMR and EAS groups, and the sample sizes for these groups were substantially smaller. Larger samples and stronger genetic instruments are needed to better assess the consistency of our findings in these ancestry groups. Fifth, we focused on genetic susceptibility, but variability in LDL-C response to diet is likely multifactorial, and additional contributors, such as metabolic health^52,53^ and food-food interactions,^11^ were not addressed.

In conclusion, genome-wide polygenic susceptibility to elevated LDL-C modifies the association between saturated fat intake and LDL-C in a nonlinear manner across the genetic spectrum. The association between saturated fat intake and LDL-C appears amplified among those with high PGS_LDL_. Secondary analyses suggested a similar pattern may also extend to incident ASCVD. These results may help to explain why individual responses to changes in diet are variable from person to person.

## Data Availability

Individual-level data from the Women's Health Initiative (WHI) are available with an approved proposal and sponsorship of a WHI investigator (https://www.whi.org/).

## Acknowledgements

The authors thank the WHI (Women’s Health Initiative) participants, clinical sites, investigators, and staff for their dedicated efforts. The WHI program is funded by the National Heart, Lung, and Blood Institute, National Institutes of Health, U.S. Department of Health and Human Services through contracts 75N92021D00001, 75N92021D00002, 75N92021D00003, 75N92021D00004, 75N92021D00005.

