## Supplementary Material for "Association between saturated fat intake and low-density lipoprotein cholesterol across the genetic spectrum: Results from the Women’s Health Initiative"

**Supplementary Table 1. Genotyping array by Women's Health Initiative (WHI) ancillary study and sample sizes included in this study.**

| WHI ancillary study | Genotyping array | Self-identified race and ethnicity | Target outcome | Sample size included in this study (%) |
|---|---|---|---|---|
| Hip Fracture (BA03) | Illumina 550K and 610K | Mostly White | Hip fracture | 190 (0.9%) |
| SHARE (M5) | Affymetrix 6.0 | Mostly Black and Hispanic | NA | 2449 (11.7%) |
| GARNET (M13) | Illumina HumanOmni1-Quad | Mostly White | Diabetes, myocardial infraction, stroke, venous thromboembolism | 3271 (15.6%) |
| WHIMS+ (W63) | Illumina HumanOmni Express Exome | Mostly White | NA | 4649 (22.2%) |
| PAGE II (AS349) | Illumina MEGA | Mostly Black and Hispanic | NA | 8542 (40.8%) |
| ONCO (M18) | Illumina OncoArray | Mostly White | Breast cancer | 1020 (4.9%) |
| LLS (W66) | Illumina HumanOmni Express Exome | Mostly White, Black and Hispanic | NA | 819 (3.9%) |
| GECCO (AS224) | Illumina 610 and Cytochip 370K | White and Black | Colorectal cancer | 0 |
| MOPMAP (AS264) | Affymetrix Gene Titan, Axiom Genome-Wide Human CEU I | White | Ventricular Ectopy | 0 |

**Supplementary Table 2. Baseline characteristics of the study cohort and the full Women's Health Initiative cohort.**

| | Study subset (genotyped participants with baseline LDL-C) (n = 20,940) | Whole WHI cohort (n = 161,808) | |
|---|---|---|---|
| | Value | Value | n missing |
| Untreated LDL-C, mg/dL | 148.0 [123.0, 175.7] | 145.4 [121.0, 173.0] | 130,173 |
| Age, years | 65.0 [58.0, 70.0] | 63.0 [57.0, 69.0] | 0 |
| Self-reported race/ethnicity [n (%)] | | | 0 |
| White | 10027 (47.9) | 133328 (82.4) | |
| Black | 6910 (33.0) | 14167 (8.8) | |
| Hispanic | 3008 (14.4) | 7312 (4.5) | |
| Other / Unidentified | 995 (4.8) | 7001 (4.3) | |
| College degree or higher [n (%)] | 6994 (33.4) | 63415 (39.5) | 1216 |
| History of ASCVD [n (%)] | 1457 (7.0) | 9722 (6.0) | 1 |
| History of diabetes [n (%)] | 1941 (9.3) | 9618 (5.9) | 110 |
| Lipid lowering drug use [n (%)] | 1891 (9.0) | 14187 (8.8) | 2 |
| Estrogen use [n (%)] | 2679 (12.8) | 51716 (32.0) | 0 |
| BMI, kg/m$^2$ | 28.6 [25.1, 32.9] | 26.9 [23.7, 31.1] | 1427 |
| Obesity (≥ 30.0 kg/m$^2$) | 8457 (40.4) | 48366 (30.2) | 1427 |
| Current smoker [n (%)] | 1887 (9.0) | 11142 (7.0) | 2126 |
| Physical activity, MET-h/week | 6.4 [1.3, 15.6] | 8.3 [2.3, 17.8] | 7471 |
| Following a low-fat diet [n (%)] | 7614 (36.4) | 65167 (41.1) | 3368 |
| Clinical trial participation [n (%)] | 15570 (74.4) | 68132 (42.1) | 0 |

Values are reported as the median [Q1, Q3] or n (%). ASCVD, atherosclerotic cardiovascular disease; BMI, body mass index; LDL-C, low-density lipoprotein cholesterol; MET-h/week, metabolic equivalent hours per week.

**Supplementary Table 3. Associations of the polygenic risk score for LDL-C ($PGS_{LDL}$) with measured LDL-C stratified by genetically inferred ancestry group among Women's Health Initiative participants.**

| Sample | n | β (mg/dL) per 1 SD [95% CI] | Incremental $R^2$ [95% CI] |
|---|---|---|---|
| European (EUR) | 10,488 | 17.0 [16.3, 17.7] | 0.178 [0.165, 0.191] |
| African (AFR) | 7,046 | 19.1 [18.1, 20.0] | 0.174 [0.156, 0.190] |
| American (AMR) | 2,996 | 15.0 [13.7, 16.4] | 0.134 [0.113, 0.158] |
| East Asian (EAS) | 410 | 10.9 [7.0, 14.9] | 0.06 [0.026, 0.115] |

Models adjust for age, $age^2$, WHI ancillary study, ancestry-specific principal components 1-10. The 95% confidence intervals for the incremental $R^2$ were estimated using the percentile bootstrap method with 5,000 bootstrap resamples.

**Supplementary Table 4. Baseline characteristics of participants included in incident ASCVD analyses by polygenic score quartiles.**

| | $PGS_{LDL}$ Q1 (n = 4211) | $PGS_{LDL}$ Q2 (n = 4210) | $PGS_{LDL}$ Q3 (n = 4207) | $PGS_{LDL}$ Q4 (n = 4207) | P trend |
|---|---|---|---|---|---|
| Untreated LDL-C, mg/dL | 127 [107, 150] | 143 [122, 167] | 154 [130, 178] | 170 [145, 197] | < 0.001 |
| Age, years | 65.0 [58.0, 70.0] | 65.0 [58.0, 70.0] | 65.0 [58.0, 70.0] | 65.0 [58.0, 70.0] | 0.91 |
| Self-reported race and ethnicity [n (%)] | | | | | 0.58 |
| Non-Hispanic White | 2088 (49.6) | 2080 (49.4) | 2109 (50.1) | 2112 (50.2) | |
| Non-Hispanic Black | 1344 (31.9) | 1303 (31.0) | 1282 (30.5) | 1251 (29.7) | |
| Hispanic | 590 (14.0) | 616 (14.6) | 622 (14.8) | 630 (15.0) | |
| Other | 189 (4.5) | 211 (5.0) | 194 (4.6) | 214 (5.1) | |
| Genetically inferred ancestry [n (%)] | | | | | 1 |
| European (EUR) | 2185 (51.9) | 2184 (51.9) | 2184 (51.9) | 2184 (51.9) | |
| African (AFR) | 1315 (31.2) | 1315 (31.2) | 1314 (31.2) | 1314 (31.2) | |
| Admixed American (AMR) | 618 (14.7) | 618 (14.7) | 617 (14.7) | 617 (14.7) | |
| East Asian (EAS) | 93 (2.2) | 93 (2.2) | 92 (2.2) | 92 (2.2) | |
| College degree or higher [n (%)] | 1450 (34.4) | 1444 (34.3) | 1390 (33.0) | 1412 (33.6) | |
| History of diabetes [n (%)] | 366 (8.7) | 352 (8.4) | 356 (8.5) | 278 (6.6) | 0.001 |
| Lipid lowering drug use [n (%)] | 153 (3.6) | 252 (6.0) | 360 (8.6) | 599 (14.2) | <0.001 |
| Estrogen use [n (%)] | 535 (12.7) | 545 (12.9) | 507 (12.1) | 544 (12.9) | 0.57 |
| BMI, $kg/m^2$ | 28.4 [25.0, 32.5] | 28.3 [24.9, 32.5] | 28.4 [24.8, 32.6] | 28.0 [24.7, 32.3] | 0.051 |
| Obesity (≥ 30.0 $kg/m^2$) | 1632 (38.8) | 1624 (38.6) | 1640 (39.0) | 1540 (36.6) | 0.093 |
| Current smoker [n (%)] | 391 (9.3) | 376 (8.9) | 397 (9.4) | 325 (7.7) | 0.024 |
| Physical activity, MET-h/week | 7.0 [1.5, 16.0] | 6.7 [1.5, 16.3] | 6.8 [1.5, 16.6] | 7.3 [1.5, 16.7] | 0.12 |
| Following a low-fat diet [n (%)] | 1321 (31.4) | 1424 (33.8) | 1601 (38.1) | 1894 (45.0) | <0.001 |
| Clinical trial participation [n (%)] | 3001 (71.3) | 2983 (70.9) | 2936 (69.8) | 2959 (70.3) | 0.48 |
| AHEI-2010 | 49.6 [42.6, 57.1] | 49.6 [42.7, 56.7] | 49.5 [42.7, 56.6] | 50.3 [43.6, 57.3] | 0.041 |

Values are reported as the median [Q1, Q3] or n (%). *P*-values compare genetic risk groups and are from Kruskal-Wallis test for continuous variables or Chi-squared tests for categorical variables. AHEI-2010, Alternative Healthy Eating Index; ASCVD, atherosclerotic cardiovascular disease; BMI, body mass index; LDL-C, low-density lipoprotein cholesterol; MET-h/week, metabolic equivalent hours per week. $PGS_{LDL}$, polygenic score for low-density lipoprotein cholesterol.

**Supplementary Table 5. Sensitivity analyses of the joint associations of genetic risk and saturated fat intake on incident ASCVD accounting for lipid-lowering therapy.**

| **(1) Excluding lipid-lowering therapy users at baseline (n = 15471)** | | | | | | |
|---|---|---|---|---|---|---|
| **All ASCVD** | | | | | | |
| Group | N | | **Model 1** | | **Model 2** | |
| | Total | Cases | HR [95% CI] | *P* | HR [95% CI] | *P* |
| $PGS_{LDL}$ Q1 – Q3 & SFA Q1 – Q3 | 7878 | 742 | Reference | - | Reference | - |
| $PGS_{LDL}$ Q1 – Q3 & SFA Q4 | 2708 | 276 | 1.02 [0.88, 1.17] | 0.83 | 1.00 [0.87, 1.16] | 0.97 |
| $PGS_{LDL}$ Q4 & SFA Q1 – Q3 | 2723 | 261 | 1.06 [0.92, 1.23] | 0.39 | 0.93 [0.80, 1.07] | 0.31 |
| $PGS_{LDL}$ Q4 & SFA Q4 | 767 | 116 | 1.47 [1.21, 1.80] | 0.0001 | 1.28 [1.05, 1.57] | 0.02 |
| Additive interaction (RERI) [95% CI] | - | - | 0.39 [0.07 0.72] | | 0.35 [0.06, 0.64] | |
| **Myocardial infarction, ischemic stroke, and coronary heart disease death** | | | | | | |
| Group | N | | **Model 1** | | **Model 2** | |
| | Total | Cases | HR [95% CI] | *P* | HR [95% CI] | *P* |
| $PGS_{LDL}$ Q1 – Q3 & SFA Q1 – Q3 | 8108 | 512 | Reference | - | Reference | - |
| $PGS_{LDL}$ Q1 – Q3 & SFA Q4 | 2796 | 188 | 0.99 [0.83, 1.18] | 0.88 | 0.98 [0.82, 1.16] | 0.82 |
| $PGS_{LDL}$ Q4 & SFA Q1 – Q3 | 2814 | 170 | 1.01 [0.85, 1.20] | 0.94 | 0.93 [0.77, 1.11] | 0.40 |
| $PGS_{LDL}$ Q4 & SFA Q4 | 797 | 86 | 1.56 [1.24, 1.97] | 0.0002 | 1.44 [1.14, 1.83] | 0.002 |
| Additive interaction (RERI) [95% CI] | - | - | 0.57 [0.17 0.96] | | 0.54 [0.17 0.90] | |
| **(2) Lipid-lowering therapy as a time-varying covariate (n = 16835)** | | | | | | |
| **All ASCVD** | | | | | | |
| Group | N | | **Model 1** | | **Model 2** | |
| | Total | Cases | HR [95% CI] | *P* | HR [95% CI] | *P* |
| $PGS_{LDL}$ Q1 – Q3 & SFA Q1 – Q3 | 8525 | 837 | Reference | - | Reference | - |
| $PGS_{LDL}$ Q1 – Q3 & SFA Q4 | 2947 | 319 | 1.04 [0.91, 1.19] | 0.56 | 1.03 [0.90, 1.17] | 0.71 |
| $PGS_{LDL}$ Q4 & SFA Q1 – Q3 | 2967 | 299 | 1.03 [0.90, 1.18] | 0.68 | 0.94 [0.82, 1.08] | 0.37 |
| $PGS_{LDL}$ Q4 & SFA Q4 | 814 | 127 | 1.44 [1.19, 1.74] | 0.0002 | 1.29 [1.06, 1.56] | 0.01 |
| Additive interaction (RERI) [95% CI] | - | - | 0.37 [0.07, 0.67] | | 0.32 [0.04, 0.60] | |
| **Myocardial infarction, ischemic stroke, and coronary heart disease death** | | | | | | |
| Group | N | | **Model 1** | | **Model 2** | |
| | Total | Cases | HR [95% CI] | *P* | HR [95% CI] | *P* |
| $PGS_{LDL}$ Q1 – Q3 & SFA Q1 – Q3 | 8794 | 568 | Reference | - | Reference | - |
| $PGS_{LDL}$ Q1 – Q3 & SFA Q4 | 3047 | 219 | 1.03 [0.88, 1.21] | 0.72 | 1.02 [0.87, 1.20] | 0.81 |
| $PGS_{LDL}$ Q4 & SFA Q1 – Q3 | 3069 | 197 | 1.03 [0.88, 1.22] | 0.71 | 0.96 [0.81, 1.14] | 0.65 |
| $PGS_{LDL}$ Q4 & SFA Q4 | 853 | 88 | 1.48 [1.18, 1.86] | 0.0008 | 1.37 [1.08, 1.72] | 0.009 |
| Additive interaction (RERI) [95% CI] | - | - | 0.42 [0.05, 0.79] | | 0.38 [0.04, 0.73] | |

Hazard ratios (HRs) and 95% CIs were estimated from Cox proportional hazards models for incident ASCVD. The RERI was calculated from the estimated HRs, and 95% CIs were obtained using the delta method. Section (1) presents analyses excluding participants using lipid-lowering therapy at baseline. Section (2) presents analyses in the full cohort with lipid-lowering therapy modeled as a time-varying covariate. Model 1 was adjusted for age, genetic principal components 1–5, ancillary study, body mass index, self-reported race and ethnicity, education, smoking, metabolic hour equivalents per week, estrogen therapy, and following a low-fat diet. Model 2 additionally adjusted for baseline untreated LDL-C levels. For both $PGS_{LDL}$ and saturated fat intake, Q4 denotes the highest quartile, and Q1-Q3 indicate the lower three quartiles. ASCVD, atherosclerotic cardiovascular disease; LDL-C, low-density lipoprotein cholesterol; $PGS_{LDL}$, polygenic score for LDL-C; RERI, relative excess risk due to interaction; SFA, saturated fat intake (% of total daily energy intake).

**Supplementary Table 6. Sensitivity analyses of the joint associations of genetic risk and saturated fat intake on incident ASCVD with an expanded set of covariates.**

| All ASCVD | | | | | | | | |
|---|---|---|---|---|---|---|---|---|
| Group | N | | **Model 1** | | **Model 2A** | | **Model 2B** | |
| | Total | Cases | HR [95% CI] | *P* | HR [95% CI] | *P* | HR [95% CI] | *P* |
| $PGS_{LDL}$ Q1-Q3 & SFA Q1-Q3 | 8521 | 837 | Reference | - | Reference | - | Reference | - |
| $PGS_{LDL}$ Q1-Q3 & SFA Q4 | 2941 | 319 | 0.99 [0.86, 1.14] | 0.89 | 0.98 [0.85, 1.12] | 0.73 | 0.98 [0.85, 1.12] | 0.72 |
| $PGS_{LDL}$ Q4 & SFA Q1-Q3 | 2965 | 299 | 1.05 [0.92, 1.20] | 0.50 | 0.95 [0.83, 1.09] | 0.40 | 0.94 [0.82, 1.08] | 0.32 |
| $PGS_{LDL}$ Q4 & SFA Q4 | 814 | 127 | 1.39 [1.15, 1.69] | 0.0007 | 1.25 [1.03, 1.52] | 0.03 | 1.24 [1.02, 1.50] | 0.03 |
| Additive interaction (RERI) [95% CI] | - | - | 0.36 [0.06, 0.65] | | 0.32 [0.04, 0.59] | | 0.32 [0.05, 0.59] | |
| **Myocardial infarction, ischemic stroke, and coronary heart disease death** | | | | | | | | |
| Group | N | | **Model 1** | | **Model 2A** | | **Model 2B** | |
| | Total | Cases | HR [95% CI] | *P* | HR [95% CI] | *P* | HR [95% CI] | *P* |
| $PGS_{LDL}$ Q1-Q3 & SFA Q1-Q3 | 8790 | 568 | Reference | - | Reference | - | Reference | - |
| $PGS_{LDL}$ Q1-Q3 & SFA Q4 | 3041 | 219 | 1.00 [0.84, 1.18] | 0.96 | 0.99 [0.84, 1.16] | 0.89 | 0.99 [0.84, 1.17] | 0.90 |
| $PGS_{LDL}$ Q4 & SFA Q1-Q3 | 3067 | 197 | 1.03 [0.87, 1.21] | 0.76 | 0.97 [0.82, 1.14] | 0.69 | 0.96 [0.81, 1.14] | 0.64 |
| $PGS_{LDL}$ Q4 & SFA Q4 | 853 | 88 | 1.43 [1.14, 1.80] | 0.002 | 1.34 [1.06, 1.69] | 0.01 | 1.33 [1.05, 1.69] | 0.02 |
| Additive interaction (RERI) [95% CI] | - | - | 0.41 [0.05, 0.77] | | 0.38 [0.04, 0.73] | | 0.38 [0.04, 0.72] | |

Hazard ratio (HR) and 95% CI from Cox proportional hazards models of incident ASCVD. The RERI was calculated from the estimated HRs, with the 95% CIs estimated using the delta method. Model 1 is adjusted for covariates included in the primary analyses with the addition of a five-category education variable, six-category income variable, four-category smoking variable, and the alternative healthy eating index (AHEI). Model 2 is a mediator-adjusted model: Model 2A adjusts for Model 1 covariates plus baseline untreated LDL-C levels and Model 2B adjusts for model 1 covariates plus baseline measured LDL-C levels. For both $PGS_{LDL}$ and saturated fat intake, Q4 denotes the highest quartile, and Q1-Q3 indicate the lower three quartiles. ASCVD, atherosclerotic cardiovascular disease; LDL-C, low density lipoprotein cholesterol; $PGS_{LDL}$, polygenic score for low-density lipoprotein cholesterol; RERI, relative excess risk due to interaction; SFA, saturated fat intake (as % of total daily calories).

**Supplementary Table 7. Sensitivity analyses of the joint associations of genetic risk and saturated fat intake on incident ASCVD using Fine-Gray competing risk models.**

| **All ASCVD** | | | | | | | | |
|---|---|---|---|---|---|---|---|---|
| Group | N | | **Model 1** | | **Model 2A** | | **Model 2B** | |
| | Total | Cases | SHR [95% CI] | *P* | SHR [95% CI] | *P* | SHR [95% CI] | *P* |
| $PGS_{LDL}$ Q1-Q3 & SFA Q1-Q3 | 8525 | 837 | Reference | - | Reference | - | Reference | - |
| $PGS_{LDL}$ Q1-Q3 & SFA Q4 | 2947 | 319 | 1.04 [0.91, 1.19] | 0.60 | 1.02 [0.89, 1.17] | 0.73 | 1.02 [0.90, 1.17] | 0.72 |
| $PGS_{LDL}$ Q4 & SFA Q1-Q3 | 2967 | 299 | 1.04 [0.91, 1.19] | 0.54 | 0.94 [0.82, 1.08] | 0.38 | 0.93 [0.81, 1.07] | 0.30 |
| $PGS_{LDL}$ Q4 & SFA Q4 | 814 | 127 | 1.43 [1.17, 1.74] | 0.0004 | 1.26 [1.03, 1.55] | 0.03 | 1.25 [1.02, 1.54] | 0.03 |
| **Myocardial infarction, ischemic stroke, and coronary heart disease death** | | | | | | | | |
| Group | N | | **Model 1** | | **Model 2A** | | **Model 2B** | |
| | Total | Cases | SHR [95% CI] | *P* | SHR [95% CI] | *P* | SHR [95% CI] | *P* |
| $PGS_{LDL}$ Q1-Q3 & SFA Q1-Q3 | 8794 | 568 | Reference | - | Reference | - | Reference | - |
| $PGS_{LDL}$ Q1-Q3 & SFA Q4 | 3047 | 219 | 1.03 [0.87, 1.21] | 0.73 | 1.02 [0.87, 1.20] | 0.80 | 1.02 [0.87, 1.20] | 0.80 |
| $PGS_{LDL}$ Q4 & SFA Q1-Q3 | 3069 | 197 | 1.02 [0.87, 1.21] | 0.80 | 0.96 [0.81, 1.14] | 0.61 | 0.95 [0.80, 1.13] | 0.55 |
| $PGS_{LDL}$ Q4 & SFA Q4 | 853 | 88 | 1.45 [1.14, 1.83] | 0.002 | 1.34 [1.05, 1.71] | 0.02 | 1.33 [1.05, 1.70] | 0.02 |

Subdistribution hazard ratio (SHR) and 95% CI from Fine-Gray competing risk models of incident ASCVD, treating non-coronary heart disease death as a competing event. Model 1 is adjusted for age, genetic principal components 1-5, ancillary study, body mass index, self-reported race and ethnicity, education, smoking, metabolic hour equivalents per week, lipid-lowering therapy, estrogen therapy, and following a low-fat diet. Model 2 is a mediator-adjusted model: Model 2A adjusts for Model 1 covariates plus baseline untreated LDL-C levels and Model 2B adjusts for model 1 covariates plus baseline measured LDL-C levels. For both $PGS_{LDL}$ and saturated fat intake, Q4 denotes the highest quartile, and Q1-Q3 indicate the lower three quartiles. ASCVD, atherosclerotic cardiovascular disease; LDL-C, low density lipoprotein cholesterol; $PGS_{LDL}$, polygenic score for low-density lipoprotein cholesterol; SFA, saturated fat intake (as % of total daily calories).

**Supplementary Table 8. Sensitivity analyses of the joint associations of genetic risk and saturated fat intake on incident ASCVD using an alternative categorization.**

| Group | N | | Model 1 | | Model 2A | | Model 2B | |
|---|---|---|---|---|---|---|---|---|
| **All ASCVD** | | | | | | | | |
| | Total | Cases | HR [95% CI] | *P* | HR [95% CI] | *P* | HR [95% CI] | *P* |
| $PGS_{LDL}$ Q1-Q3 & SFA < median | 5583 | 528 | Reference | - | Reference | - | Reference | - |
| $PGS_{LDL}$ Q1-Q3 & SFA ≥ median | 5889 | 628 | 1.08 [0.95, 1.22] | 0.22 | 1.07 [0.95, 1.21] | 0.27 | 1.07 [0.95, 1.22] | 0.26 |
| $PGS_{LDL}$ Q4 & SFA < median | 2111 | 195 | 0.98 [0.83, 1.16] | 0.85 | 0.89 [0.75, 1.06] | 0.19 | 0.88 [0.75, 1.05] | 0.15 |
| $PGS_{LDL}$ Q4 & SFA ≥ median | 1670 | 231 | 1.39 [1.19, 1.63] | 4.7e-5 | 1.24 [1.06, 1.46] | 0.009 | 1.23 [1.05, 1.45] | 0.01 |
| Additive interaction (RERI) [95% CI] | - | - | 0.33 [0.08, 0.58] | | 0.28 [0.05, 0.51] | | 0.27 [0.05, 0.50] | |
| **Myocardial infarction, ischemic stroke, and coronary heart disease death** | | | | | | | | |
| Group | N | | **Model 1** | | **Model 2A** | | **Model 2B** | |
| | Total | Cases | HR [95% CI] | *P* | HR [95% CI] | *P* | HR [95% CI] | *P* |
| $PGS_{LDL}$ Q1-Q3 & SFA < median | 5763 | 348 | Reference | - | Reference | - | Reference | - |
| $PGS_{LDL}$ Q1-Q3 & SFA ≥ median | 6078 | 439 | 1.12 [0.96, 1.30] | 0.96 | 1.11 [0.96, 1.29] | 0.16 | 1.11 [0.96, 1.30] | 0.16 |
| $PGS_{LDL}$ Q4 & SFA < median | 2179 | 127 | 0.98 [0.80, 1.21] | 0.76 | 0.92 [0.75, 1.14] | 0.45 | 0.92 [0.75, 1.13] | 0.42 |
| $PGS_{LDL}$ Q4 & SFA ≥ median | 1743 | 158 | 1.42 [1.17, 1.72] | 0.0004 | 1.32 [1.08, 1.61] | 0.006 | 1.32 [1.08, 1.60] | 0.007 |
| Additive interaction (RERI) [95% CI] | - | - | 0.32 [0.009, 0.62] | | 0.29 [-0.007, 0.58] | | 0.28 [-0.007, 0.57] | |

Hazard ratio (HR) and 95% CI from Cox proportional hazards models of incident ASCVD. The RERI was calculated from the estimated HRs, with the 95% CIs estimated using the delta method. Model 1 is adjusted for age, genetic principal components 1-5, ancillary study, body mass index, self-reported race and ethnicity, education, smoking, metabolic hour equivalents per week, lipid-lowering therapy, estrogen therapy, and following a low-fat diet. Model 2 is a mediator-adjusted model: Model 2A adjusts for Model 1 covariates plus baseline untreated LDL-C levels and Model 2B adjusts for model 1 covariates plus baseline measured LDL-C levels. For $PGS_{LDL}$, Q4 denotes the highest quartile, and Q1-Q3 indicate the lower three quartiles. SFA is split at the median. ASCVD, atherosclerotic cardiovascular disease; LDL-C, low density lipoprotein cholesterol; $PGS_{LDL}$, polygenic score for low-density lipoprotein cholesterol; RERI, relative excess risk due to interaction; SFA, saturated fat intake (as % of total daily calories).

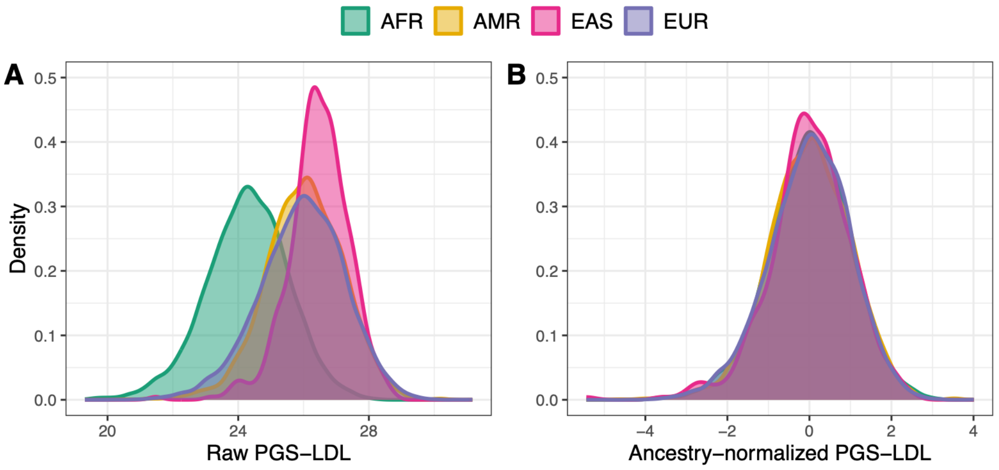


**Supplementary Figure 1. Distributions of the raw and ancestry-normalized polygenic risk score for LDL-C stratified by ancestry group among Women's Health Initiative participants.** AFR, African ancestry (n = 7,046); AMR, Admixed American ancestry AMR (n = 2,996); EAS, East Asian ancestry (n = 410); EUR, European ancestry (n = 10,488); PGS-LDL, polygenic risk score for low-density lipoprotein cholesterol.

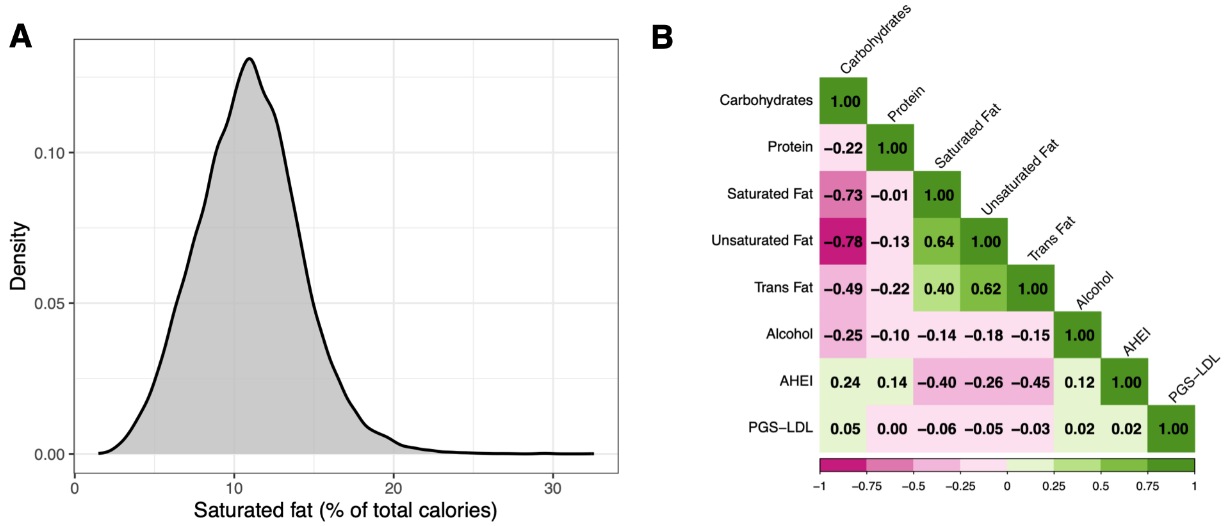


**Supplementary Figure 2. Distribution of saturated fat intake (A) and pairwise correlations between macronutrients, the diet quality score, and the polygenic score for LDL-C (B) among Women's Health Initiative participants.** The correlation between $PGS_{LDL}$ and saturated fat intake (% total calories) was similar across ancestry groups (EUR r = -0.07; AFR r = -0.05; AMR r = -0.07; EAS r = -0.04). AHEI, Alternative Healthy Eating Index; PGS-LDL, polygenic score for low-density lipoprotein cholesterol.

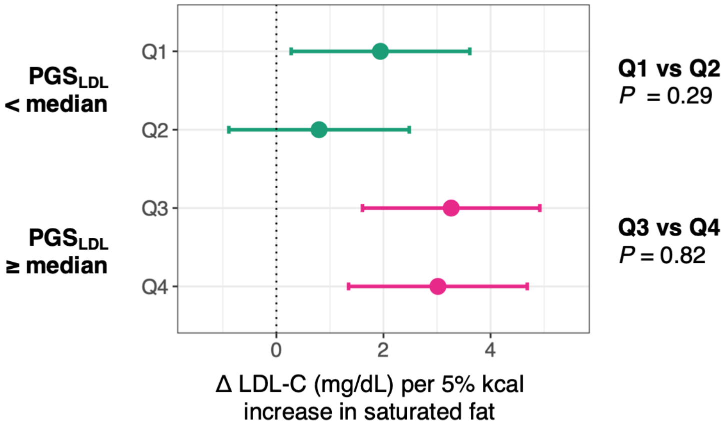


**Supplementary Figure 3. Association of saturated fat intake with LDL-C by polygenic score quartiles.** Forest plot shows the β (95% CI) representing the estimated change in LDL-C (mg/dL) associated with each 5% of total energy increase in saturated fat in place of carbohydrates, stratified by $PGS_{LDL}$ quartile. *P*-values on the right comparing adjacent $PGS_{LDL}$ quartiles were obtained from pairwise contrasts estimated using the *emmeans* R package. $PGS_{LDL}$, polygenic score for low-density lipoprotein cholesterol.

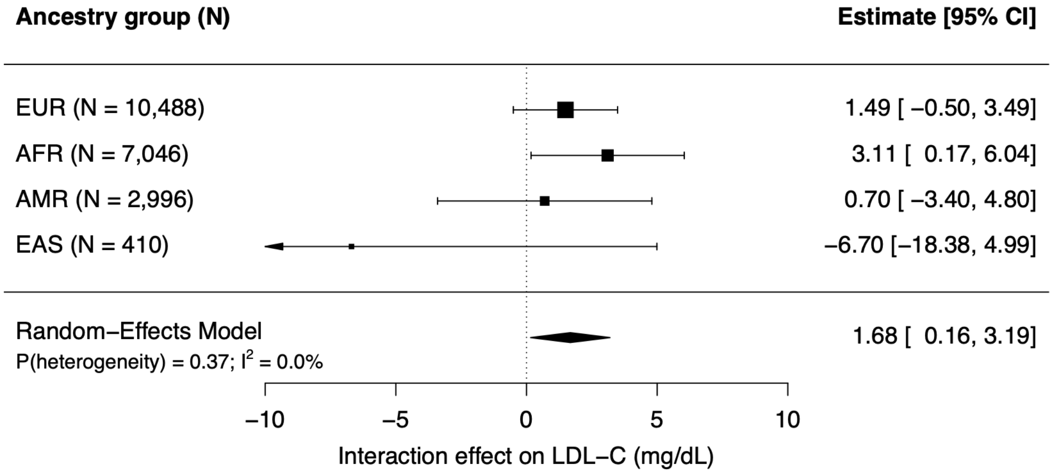


**Supplementary Figure 4. Interaction effect of polygenic score and saturated fat intake on LDL-C stratified by genetic ancestry group, with random-effects meta-analysis.** Squares represent ancestry-specific interaction coefficients (β) and 95% CIs, and the diamond represents the pooled random-effects meta-analysis estimate. The interaction reflects the difference in the association between 5% kcal from saturated fat (in place on carbohydrates) and LDL-C among participants with $PGS_{LDL}$ above versus below the median (reference group). Models adjust for age, $age^2$, ancestry-specific genetic principal components 1-5, ancillary study, body mass index, self-reported race and ethnicity, education, smoking, metabolic hour equivalents per week, history of atherosclerotic cardiovascular disease, history of diabetes, lipid lowering therapy, estrogen therapy, and following a low-fat diet. AFR, African ancestry; AMR, Admixed American ancestry; EAS, East Asian ancestry; EUR, European ancestry; $PGS_{LDL}$, polygenic score for low-density lipoprotein cholesterol.

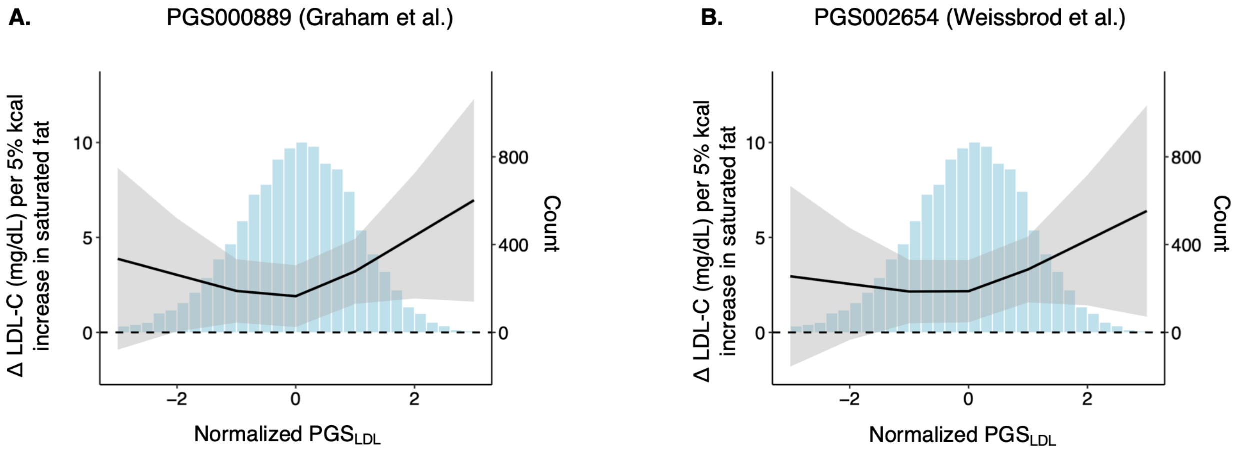


**Supplementary Figure 5. Restricted cubic spline plots of the association between saturated fat intake and LDL-C across the genetic risk spectrum, summarized using (A) the primary multi-ancestry $PGS_{LDL}$ and (B) an alternative European-derived $PGS_{LDL}$ among WHI participants of European ancestry (n = 10,488).** Splines with three knots model the effect of saturated fat intake (in place of carbohydrates) on LDL-C (left y-axis) across the genetic risk spectrum (x-axis). The histogram is referenced to the right y-axis and depicts the distribution of participants across the $PGS_{LDL}$ spectrum. Models adjust for age, $age^2$, genetic principal components 1-5, ancillary study, body mass index, self-reported race and ethnicity, education, smoking, metabolic hour equivalents per week, history of atherosclerotic cardiovascular disease, history of diabetes, lipid lowering therapy, estrogen therapy, and following a low-fat diet. LDL-C, low-density lipoprotein cholesterol; $PGS_{LDL}$, polygenic score for LDL-C.

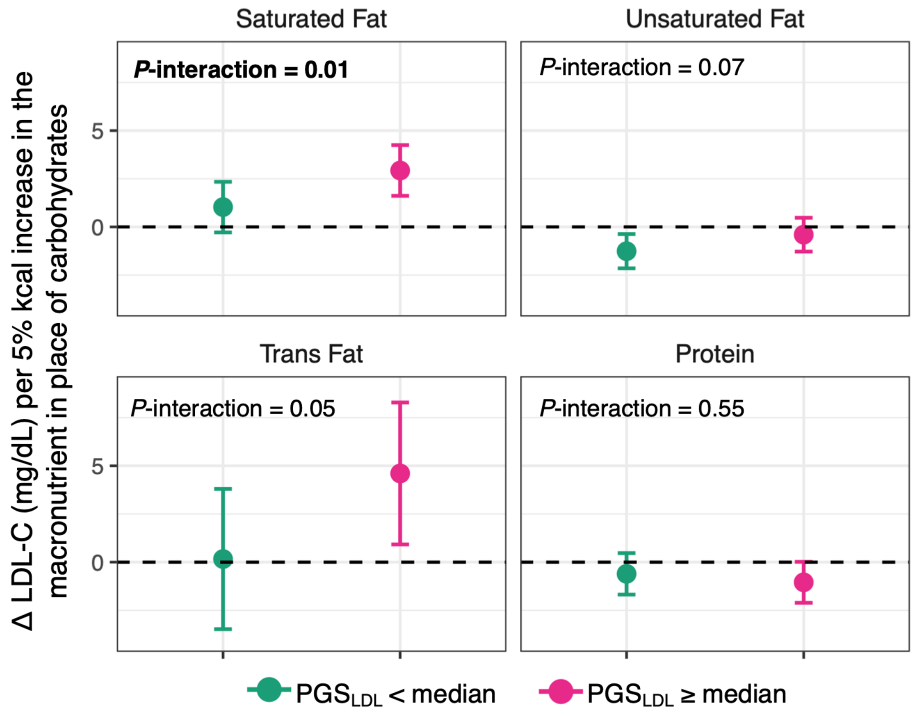


**Supplementary Figure 6. Assessment for interaction between polygenic score and other macronutrients in the substitution model.** Forest plots show the β (95%CIs) representing the change in LDL-C (mg/dL) per 5% of total calories increase in each macronutrient in place of carbohydrates, stratified by $PGS_{LDL}$ category. All models adjust for age, age$^2$, genetic principal components 1-5, ancillary study, body mass index, self-reported race and ethnicity, education, smoking, metabolic hour equivalents per week, history of atherosclerotic cardiovascular disease, history of diabetes, lipid lowering therapy, estrogen therapy, and following a low-fat diet. $PGS_{LDL}$, polygenic score for low-density lipoprotein cholesterol.

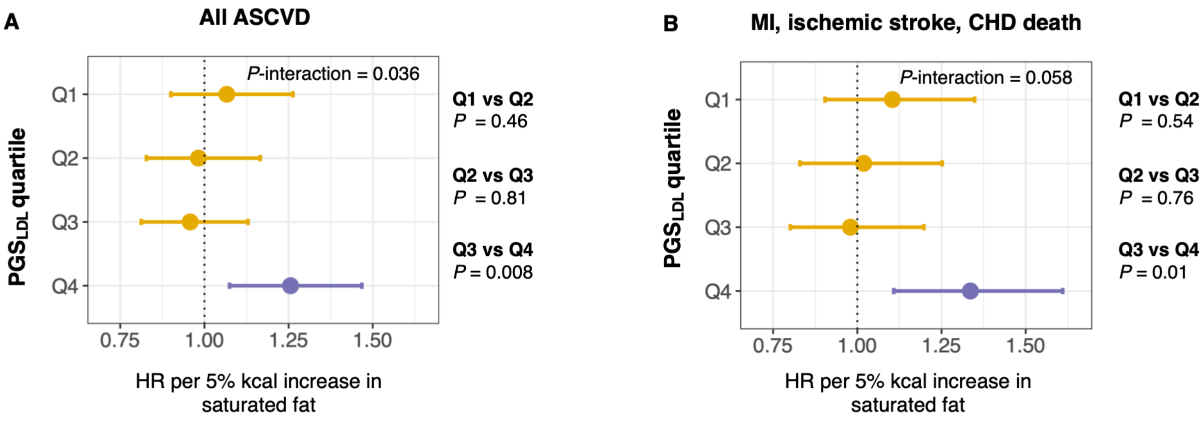


**Supplementary Figure 7. Association of saturated fat intake with incident ASCVD by polygenic score quartile.** Forest plots show the hazard ratio (HR) and 95% CI for (A) all ASCVD and (B) the strict composite of myocardial infarction, ischemic stroke, and coronary heart disease death, associated with each 5% of total energy increase in saturated fat in place of carbohydrates by $PGS_{LDL}$ quartile. *P*-interaction values were obtained from likelihood ratio tests comparing models with and without the saturated fat x $PGS_{LDL}$ interaction term. *P*-values on the right comparing adjacent $PGS_{LDL}$ quartiles were obtained from pairwise contrasts estimated using the *emmeans* R package. Models are adjusted for age, genetic principal components 1-5, ancillary study, body mass index, self-reported race and ethnicity, education, smoking, metabolic hour equivalents per week, history of diabetes, lipid-lowering therapy, estrogen therapy, and following a low-fat diet. ASCVD, atherosclerotic cardiovascular disease; $PGS_{LDL}$, polygenic score for low-density lipoprotein cholesterol.

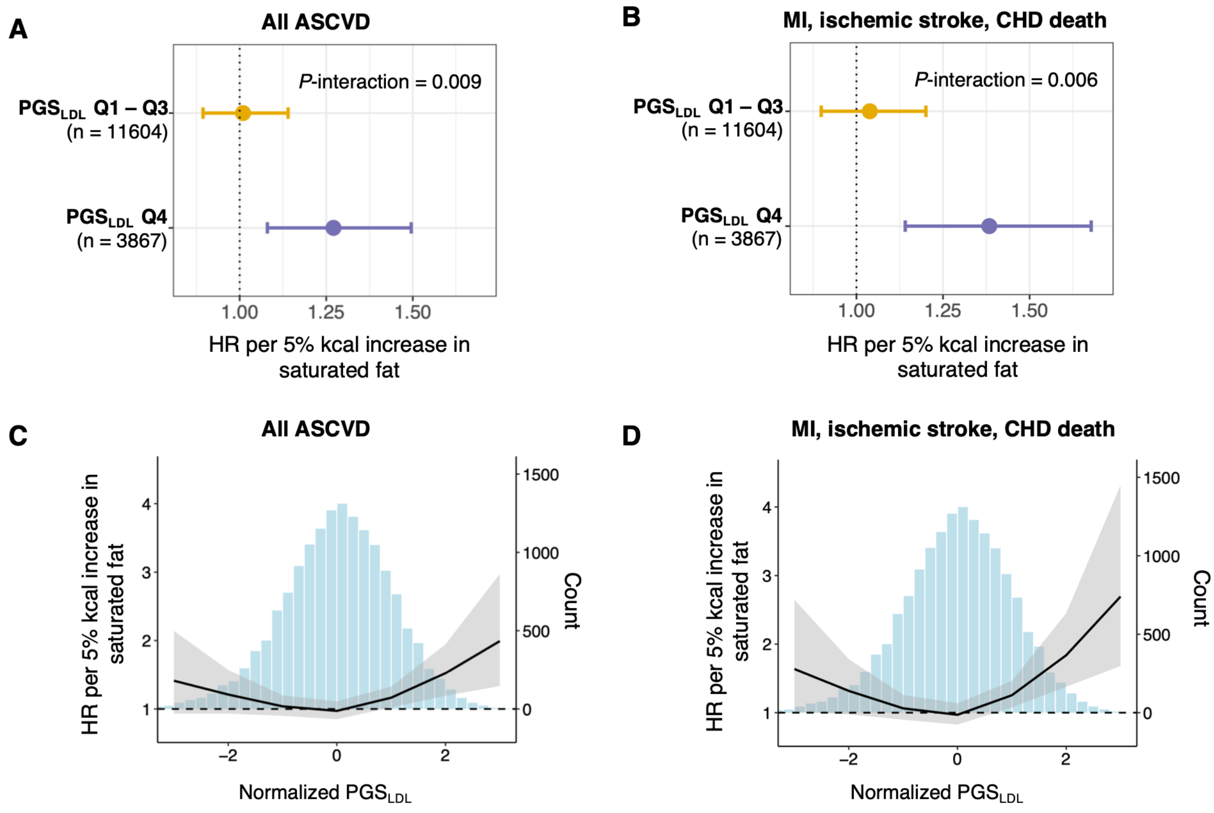


**Supplementary Figure 8. Association of saturated fat intake with incident ASCVD according to polygenic score among participants not using lipid-lowering therapy at baseline (n = 15,471).** Forest plots show the hazard ratio (HR) and 95% CI for (A) all ASCVD and (B) the strict composite of myocardial infarction, ischemic stroke, and coronary heart disease death, associated with each 5% of total energy increase in saturated fat in place of carbohydrates by $PGS_{LDL}$ category; Q4 indicates the highest quartile of $PGS_{LDL}$, and Q1-Q3 indicate the lower three quartiles. Restricted cubic spline plots show the estimated HR and 95% CI for (C) all ASCVD or (D) the strict composite of myocardial infarction, ischemic stroke, and coronary heart disease death, associated with each 5% of total energy increase in saturated fat in place of carbohydrates (left y-axis) across the $PGS_{LDL}$ spectrum (x-axis). The histogram in panels C and D, referenced to the right y-axis, depicts the distribution of participants across the $PGS_{LDL}$ spectrum. All analyses are adjusted for age, genetic principal components 1-5, ancillary study, body mass index, self-reported race and ethnicity, education, smoking, metabolic hour equivalents per week, history of diabetes, estrogen therapy, and following a low-fat diet. ASCVD, atherosclerotic cardiovascular disease; $PGS_{LDL}$, polygenic score for low-density lipoprotein cholesterol.

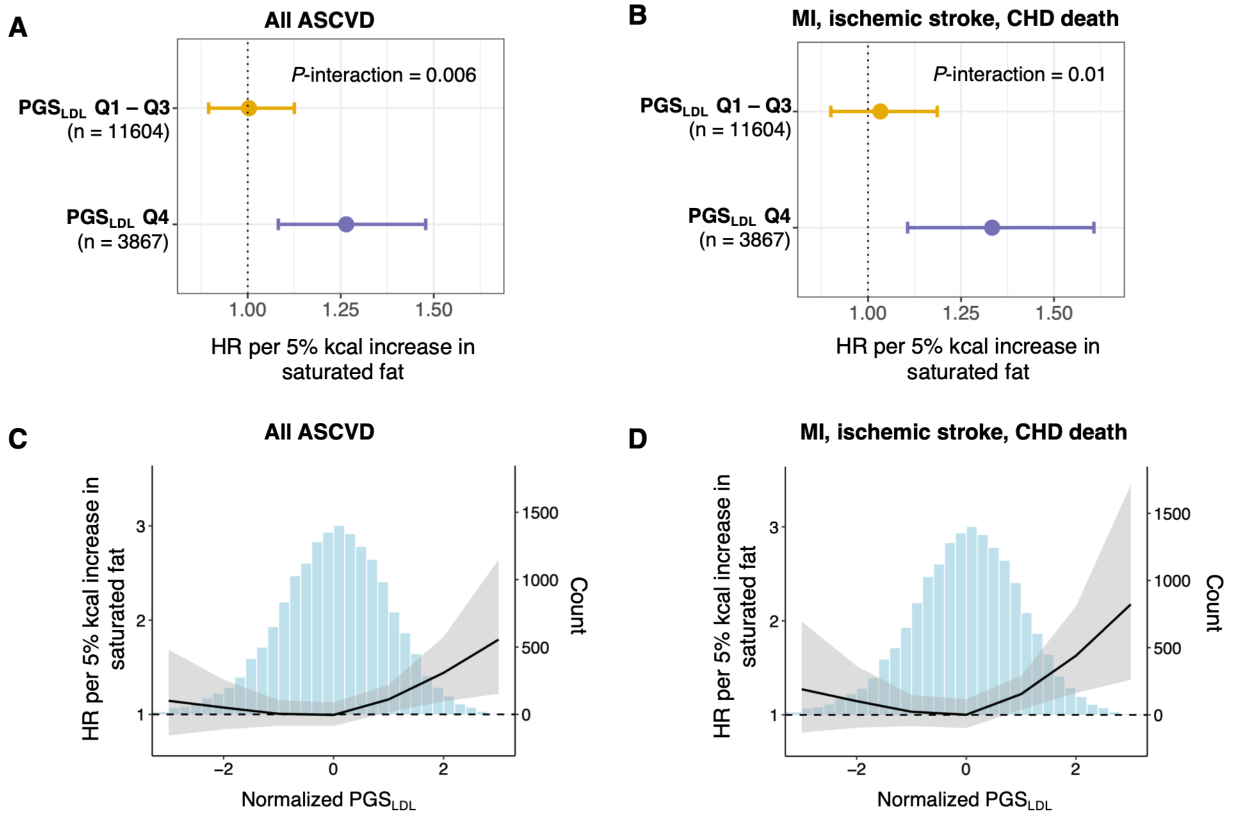


**Supplementary Figure 9. Association of saturated fat intake with incident ASCVD according to polygenic score in models incorporating lipid-lowering therapy as a time-varying covariate.** Forest plots show the hazard ratio (HR) and 95% CI for (A) all ASCVD and (B) the strict composite of myocardial infarction, ischemic stroke, and coronary heart disease death, associated with each 5% of total energy increase in saturated fat in place of carbohydrates by $PGS_{LDL}$ category; Q4 indicates the highest quartile of $PGS_{LDL}$, and Q1-Q3 indicate the lower three quartiles. Restricted cubic spline plots show the estimated HR and 95% CI for (C) all ASCVD or (D) the strict composite of myocardial infarction, ischemic stroke, and coronary heart disease death, associated with each 5% of total energy increase in saturated fat in place of carbohydrates (left y-axis) across the $PGS_{LDL}$ spectrum (x-axis). The histogram in panels C and D, referenced to the right y-axis, depicts the distribution of participants across the $PGS_{LDL}$ spectrum. The expanded set of covariates include a five-category education variable, a six-category income variable, and four-category smoking variable in addition to age, genetic principal components 1-5, ancillary study, body mass index, self-reported race and ethnicity, metabolic hour equivalents per week, history of diabetes, estrogen therapy, and following a low-fat diet. ASCVD, atherosclerotic cardiovascular disease; $PGS_{LDL}$, polygenic score for low-density lipoprotein cholesterol.

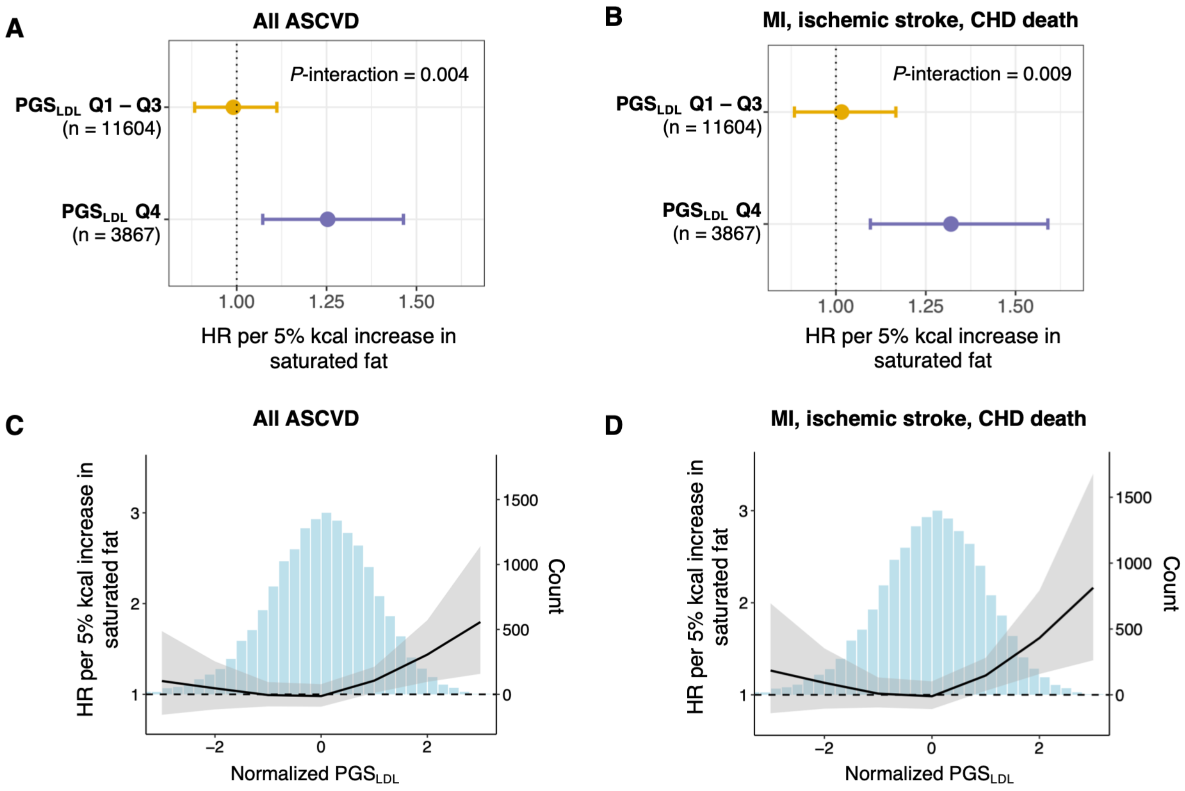


**Supplementary Figure 10. Association of saturated fat intake with incident ASCVD according to polygenic score in models with additional adjustment for an expanded set of covariates.** Forest plots show the hazard ratio (HR) and 95% CI for (A) all ASCVD and (B) the strict composite of myocardial infarction, ischemic stroke, and coronary heart disease death, associated with each 5% of total energy increase in saturated fat in place of carbohydrates by $PGS_{LDL}$ category; Q4 indicates the highest quartile of $PGS_{LDL}$, and Q1-Q3 indicate the lower three quartiles. Restricted cubic spline plots show the estimated HR and 95% CI for (C) all ASCVD or (D) the strict composite of myocardial infarction, ischemic stroke, and coronary heart disease death, associated with each 5% of total energy increase in saturated fat in place of carbohydrates (left y-axis) across the $PGS_{LDL}$ spectrum (x-axis). The histogram in panels C and D, referenced to the right y-axis, depicts the distribution of participants across the $PGS_{LDL}$ spectrum. All analyses are adjusted for covariates included in the primary analyses with the addition of a five-category education variable, six-category income variable, and four-category smoking status and pack-year variable. ASCVD, atherosclerotic cardiovascular disease; $PGS_{LDL}$, polygenic score for low-density lipoprotein cholesterol.

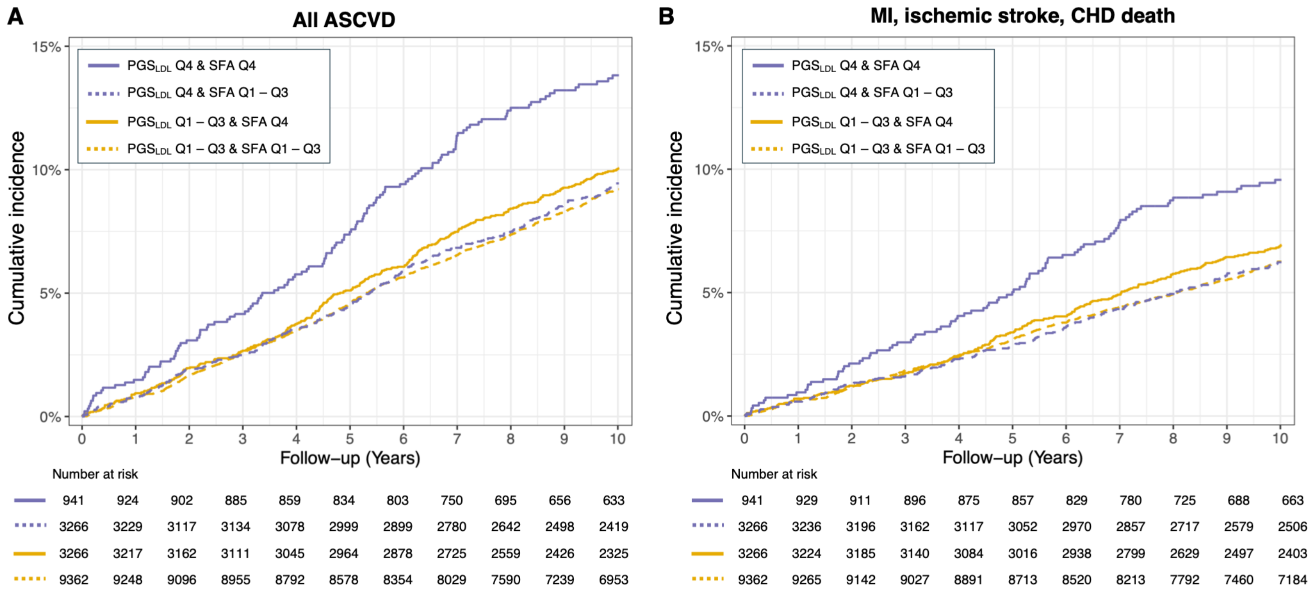


**Supplementary Figure 11. Cumulative incidence functions (competing-risk analysis) for (A) all ASCVD and (B) the strict composite of myocardial infarction, ischemic stroke, and coronary heart disease death, according to joint categories of polygenic score and saturated fat intake.** For both $PGS_{LDL}$ and saturated fat intake, Q4 denotes the highest quartile, and Q1-Q3 indicate the lower three quartiles. ASCVD, atherosclerotic cardiovascular disease; $PGS_{LDL}$, polygenic score for low-density lipoprotein cholesterol; SFA, saturated fat intake (as % of total daily energy intake).
